# An Interpretable Machine Learning Tool for In-Home Screening of Agitation Episodes in People Living with Dementia

**DOI:** 10.1101/2024.07.30.24311178

**Authors:** Marirena Bafaloukou, Ann-Kathrin Schalkamp, Nan Fletcher-Lloyd, Alex Capstick, Chloe Walsh, Cynthia Sandor, Samaneh Kouchaki, CR&T Group, Ramin Nilforooshan, Payam Barnaghi

## Abstract

**Background:** Agitation affects around 30% of people living with dementia (PLwD), increasing carer burden and straining care services. Agitation screening typically relies on subjective clinical scales and direct patient observation, which are resource-intensive and challenging to incorporate into routine care. Clinical applicability of data-driven methods for agitation screening is limited by constraints such as short observational periods, data granularity, and lack of interpretability and generalisability. Current interventions for agitation are primarily medication-based, which may lead to severe side effects and lack personalisation. Understanding how real-world factors affect agitation within home settings offers a promising avenue towards identifying potential personalised non-pharmacological interventions.

**Methods:** We used longitudinal data (32,896 person-days from n=63 PLwD) collected using in-home monitoring devices. Employing machine learning techniques, we developed a screening tool to determine the weekly risk of agitation. We incorporated a traffic-light system for risk stratification to aid clinical decision-making and employed the SHapley Additive exPlanations (SHAP) framework to increase interpretability. We designed an interactive tool that enables the exploration of personalised non-pharmacological interventions, such as modifying ambient light and temperature.

**Results:** Light Gradient-boosting Machine (LightGBM) achieved the highest performance in identifying agitation with a sensitivity of 71.32*±*7.38% and specificity of 75.28%. Implementing the traffic-light system for risk stratification increased specificity by 15% and improved all metrics. Significant contributors to agitation included low nocturnal respiratory rate, heightened alertness during sleep, and increased indoor illuminance, as revealed by statistical and feature importance analysis. Using our interactive tool, we identified that adjusting indoor lighting levels and temperature were promising and feasible interventions within our cohort.

**Conclusions:** Our interpretable framework for agitation screening, developed using data from a dementia care study, showcases significant clinical value. The accompanying interactive interface allows for the *in-silico* simulation of non-pharmacological interventions, facilitating the design of personalised interventions that can improve in-home dementia care.

## Introduction

Agitation is a neuropsychiatric symptom that affects more than 30% of people living with dementia (PLwD)^1^, and is characterised by a 93% recursion rate^2^. It is defined by inappropriate verbal, vocal, or motor actions, unrelated to the immediate needs of the individual^3^. Agitation episodes can range from non-aggressive behaviours, such as wandering and repetition, to aggressive actions^4,5^. Such episodes are associated with adverse healthcare events, including fall-related injuries and undiagnosed infections^6^. Agitation places a significant burden on carers and contributes to their sustained stress levels^7^. Identification and treatment of agitation are essential for mitigating associated risks, lowering healthcare expenses, and alleviating carer strain^6^.

Various clinical measures have been developed to assess agitation, which can be grouped into two main categories: informant ratings, including the Cohen-Mansfield Agitation Inventory (CMA-I)^4^, and direct observational assessments such as the Pittsburgh Agitation Scale^8^. Informant ratings, as noted by Cohen-Mansfield *et al.*, can be unreliable due to the personal bias of the informant, memory inaccuracies, and stress. Direct observational assessments are more objective; however, they require trained staff to monitor behaviour, making these assessments resource-intensive and not applicable to at-home settings^9,10^.

Applying machine learning (ML) methods to continuously collected remote monitoring data can address the above-mentioned limitations^11,12^. Khan *et al.*, proposed a model that detects agitation through motion and physiological sensor data using a 1-minute time-window. Their study’s small cohort (n=2) and lack of longitudinal data (28 days) limit the reliability of their findings. Moreover, the small data granularity (1-minute) restricts the exploration of long-term agitation-driving factors, thereby impacting the model’s clinical applicability. In our previous work, we integrated activity and physiological data - from 46 PLwD, collected over 2 years - into a deep learning (DL) model to predict agitation using a 6-hour window^12^. The model’s high false positive rate and the complexity of the DL model, providing limited insights into the underlying factors, hindered its clinical utility. Hekmatiathar *et al.,* incorporated interior ambient data collected over 64 days, from the living environment of one participant in a DL model, to forecast agitation using a rolling window of 30 minutes^13^. Their approach relied on frequent validation, thereby increasing the likelihood of false positives, which could disrupt patients’ and carers’ routines. Moreover, their reliance solely on environmental data and validation on only one participant limit the model’s clinical reliability. Overall, existing studies on the identification of agitation lack generalisability due to small data sizes, while studies on agitation prediction lack clinical and care applicability, as agitation antecedents are often unpredictable^14^. This highlights the need to shift towards agitation screening and develop transparent and interpretable ML models that encompass a diverse range of variables and can capture broader agitation patterns.

The National Institute of Health and Care Excellence (NICE) in the UK recommends prioritising non-pharmacological interventions, such as sensory stimulation, as the primary strategy to address dementia-related agitation^15^. This is driven by concerns surrounding potential side effects linked to medications and their limited long-term efficacy^16^. Light-exposure^17^ and music-related interventions have shown promise for the treatment of agitation in dementia^18^. However, existing studies for non-pharmacological interventions are limited by small sample sizes and inadequate reporting of findings, preventing a comprehensive understanding of non-pharmacological interventions’ efficacy^18^. Optimal design of such interventions requires a personalised study of individual patterns, symptoms and preferences^19^, an area which remains under-explored.

Here, we used real-world data from the observational clinical study called Minder^20^ to develop an ML framework that estimates and analyses the risk of agitation occurrence within a week. Using the passive monitoring data from Minder, we integrated a diverse array of features, including sleep measures, activity levels, and environmental parameters, into our model. Our dataset comprised data collected over 32,896 person-days from 63 PLwD, of which 4,096 were labelled for agitation occurrence. Through the assessment of feature importance, we sought to provide information on the model’s decision-making process. We developed a novel interactive tool, tailored for designing and validating non-pharmacological interventions, based on real-world insights from personalised agitation driving factors.

## Results

### Agitation episodes in Minder

Agitation status was determined through responses from a weekly clinical monitoring process conducted in the Minder study. Most participants lived with carers or study partners (Table S1). During the weekly monitoring process, study partners and/or participants were contacted to report the presence or absence of participants’ agitation symptoms regarding the preceding week. For our analysis, the whole preceding week, including the day of the reporting itself (8 days), was labelled as a binary target variable representing the presence or absence of agitation (see Data Collection section).

Our approach (see Figure 1) included labelled data collected in the Minder study between July 15, 2021, and March 16, 2023. For the development of our machine learning model, we focused on individuals with verified episodes of agitation or absence thereof, resulting in a final sub-cohort of 63 participants (41 males and 22 females). This sub-cohort had a total of 242 weeks labelled as positive for agitation and 270 weeks labelled as negative S1.

**Figure 1:**
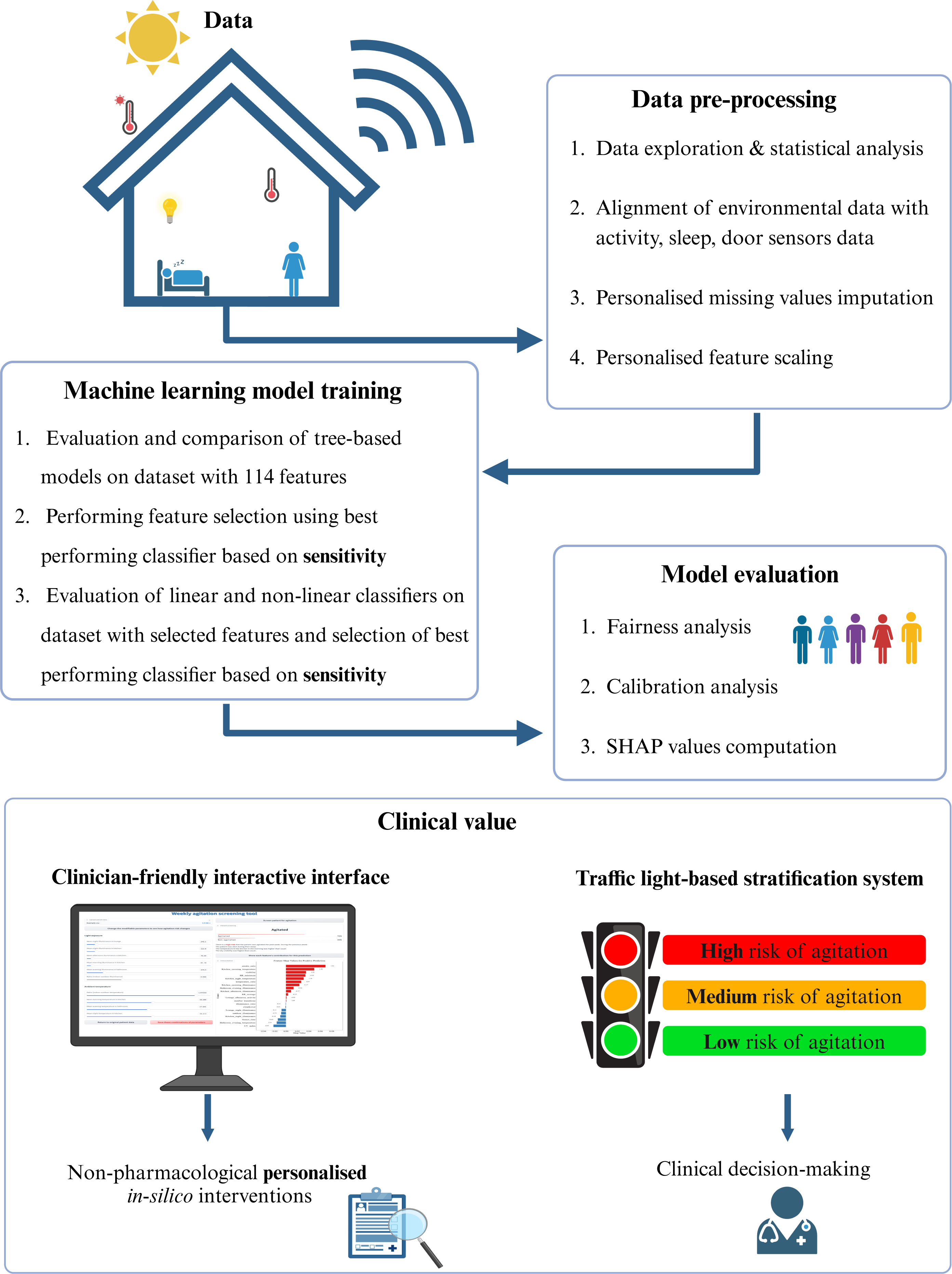
Overview of approach. The data processing and analysis pipeline are shown, illustrating the specific steps followed. Our two strategies for enhanced clinical value are showcased: an interactive interface for personalised in-silico intervention experiments and a traffic-light system to minimise false alerts. Figure was created with Biorender.

### Agitation episodes are accompanied by poor sleep quality and differences in light exposure

We conducted two-sided paired Student’s T-tests to compare extracted measures such as sleep, illuminance, and ambient temperature from the Minder in-home monitoring sensors and the Visual Crossing weather API between agitated and non-agitated weeks^21^. For the statistical analysis, we only included participants who had both positive and negative agitation events (n=29) to allow for a paired comparison (Table S1).

Sleep, illuminance and temperature features, derived from the in-home PIR sensors and the sleep mat, alongside outdoor ambient data, highlight differences between agitation and non-agitation weeks, showing their potential to be used for agitation screening. During agitation weeks, the average awake ratio was significantly higher compared to non-agitation weeks (0.27 ± 0.67 vs. −0.18 ± 0.40) (*p − value* = 9 *×* 10*^−^*^3^), whereas the average minimum respiratory rate was significantly lower (−0.33 ± 0.68 vs. 0.09 ± 0.33) (*p − value* = 2 *×* 10*^−^*^2^) (Table S14). Furthermore, indoor illuminance was significantly higher during agitation weeks compared to non-agitation weeks (0.10 ± 0.25 vs. −0.05 ± 0.19) (*p−value* = 4 *×* 10*^−^*^2^), while the indoor to outdoor illuminance (illuminance ratio) was significantly lower (−0.26 ± 0.27 vs. 0.18 ± 0.26) (*p−value <* 10*^−^*^3^) (Table S16, S21).

This feature analysis showed that sleep patterns and environmental factors differed during agitation weeks, and could be used in an ML model for screening of agitation episodes.

### Light Gradient-Boosting Machine classifier can identify agitation using sensor data

We derived 114 variables, including sleep, activity, indoor and outdoor light exposure, indoor and outdoor temperature, and seasonality, to be used in our machine learning pipeline to identify agitation(Table S3). Feature selection was performed using the SHapley Additive exPlanation (SHAP)^22^ values, computed by a Random Forest Classifier (RF), using 10 folds. This resulted in 20 features that were used for agitation screening. The investigated models include SVM, LR, LightGBM, RF, XGBoost, ADABoost, and an MLP. Model evaluation focused on maximising sensitivity, ensuring the model’s effectiveness as a sensitive screening tool.

The LightGBM yielded significantly higher sensitivity than logistic regression (*p − value < ×*10*^−^*^3^) and the other examined tree-based models (*p − value <* 5 *×* 10*^−^*^3^) (see Table S6). The LightGBM classifier also achieved higher scores in most performance metrics compared to the examined models (see Table S5). The model showed robust results in calibration (Figure S3), reliability (Figure S4), and bias analysis (Table S8, Table S9, Figure S2).

We enhanced our model’s clinical value by implementing a traffic-light risk stratification system, categorising agitation predictions into high (Red), moderate (Amber), and low (Green) risk groups. Incorporating a risk stratification via a traffic-light system increased sensitivity and specificity by 15% and improved all metrics. The inclusion of the Amber group reduced the false alerts, increasing the model’s clinical applicability. Refer to Table 1 for a comparison of metrics before and after risk stratification, and see Figure S7 for visual representations of the receiver operating characteristic (ROC) and precision-recall (PR) curves. More details on stratification are provided in section S1.4.3.

**Table 1:** Performance comparison before and after risk stratification. The traffic-light system optimised the thresholds for decision-making, creating Green (low agitation risk), Amber (medium agitation risk), and Red (high agitation risk) groups. The performance metrics from the 10-Fold cross-validation are reported as mean ± standard deviation. Metrics include accuracy, F1-score, precision, sensitivity, specificity, PR AUC (area under the precision-recall curve), ROC AUC (area under the receiver operating characteristic curve)

| Metric | Binary classification | Traffic-light system |
| --- | --- | --- |
| Accuracy | 71.36 ± 7.21 | 78.87 ± 6.88 |
| F1 Score | 71.12 ± 7.28 | 76.70 ± 7.52 |
| Precision | 71.81 ± 7.94 | 80.06 ± 7.73 |
| Sensitivity | 71.32 ± 7.38 | 76.36 ± 7.45 |
| Specificity | 75.28 ± 10.43 | 90.33 ± 7.55 |
| PR AUC | 78.60 ± 7.57 | 81.42 ± 9.99 |
| ROC AUC | 77.63 ± 6.59 | 80.73 ± 9.37 |

### Low respiratory rate and high indoor artificial illuminance contribute the most to agitation risk

To increase the clinical value of the presented model, we investigated feature importance via SHAP values.

Figure 2 demonstrates the contribution of each feature’s high (red) and low (blue) values to the model prediction. The most important features for agitation identification were low respiratory rate, high awake ratio, extreme values of visibility and indoor illuminance (both high and low), low illuminance ratio, high temperature ratio and low indoor temperatures.

**Figure 2:**
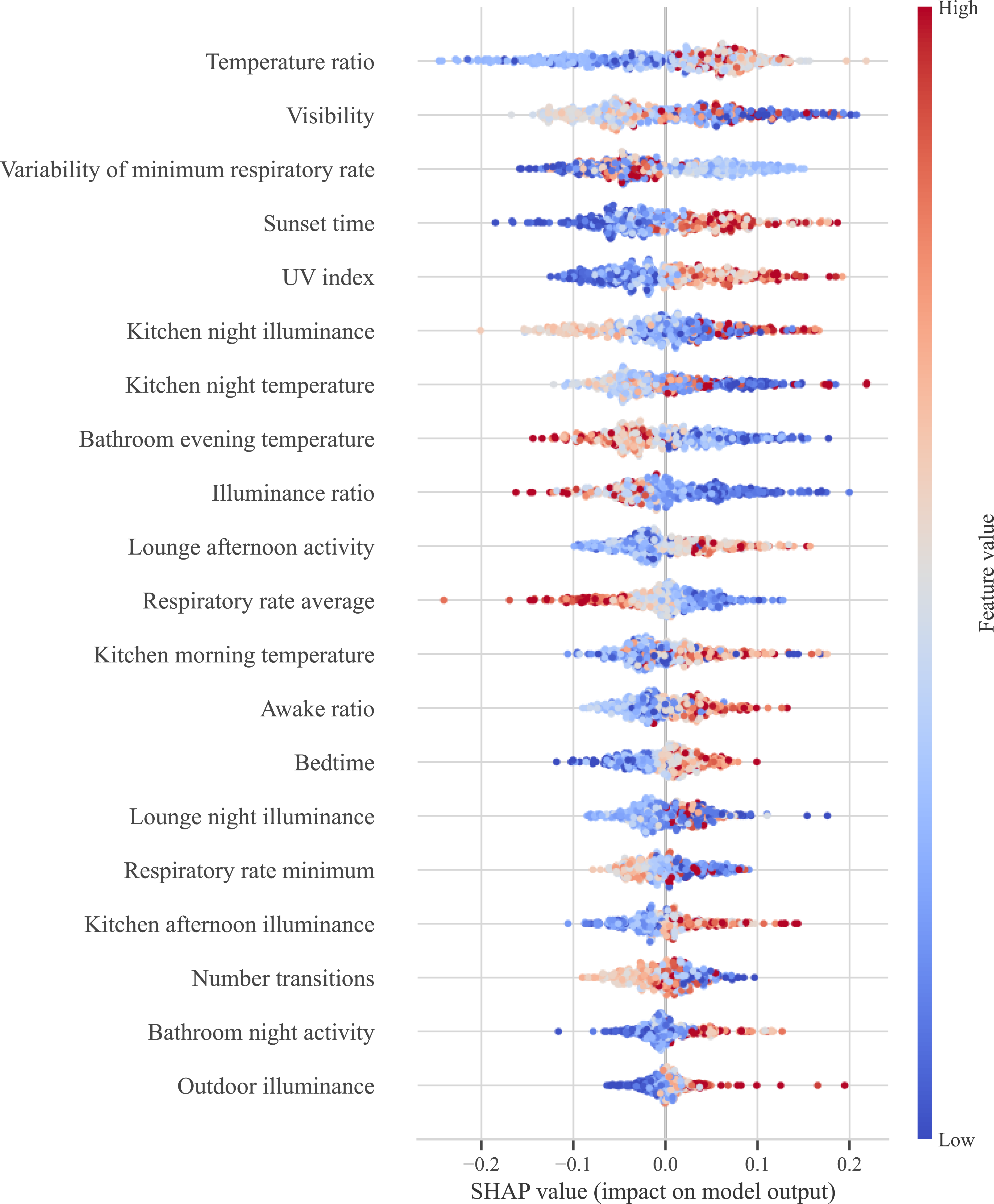
Model interpretability through feature importance analysis. The feature importance calculated using SHAP on the test sets from the 10-Fold cross-validation is shown in a summary plot. The colour represents the scaled feature value (red corresponding to higher values, blue to lower). The position of the x-axis represents the contribution of each normalised feature value to the positive prediction of agitation.

### Interactive online tool allows simulation of personalised non-pharmacological interventions

The SHAP framework allows further analysis of the features contributing to an individual prediction. Figure 3 shows two examples of one positive and one negative event from the same PLwD. Such insights can identify personalised interventions.

**Figure 3:**
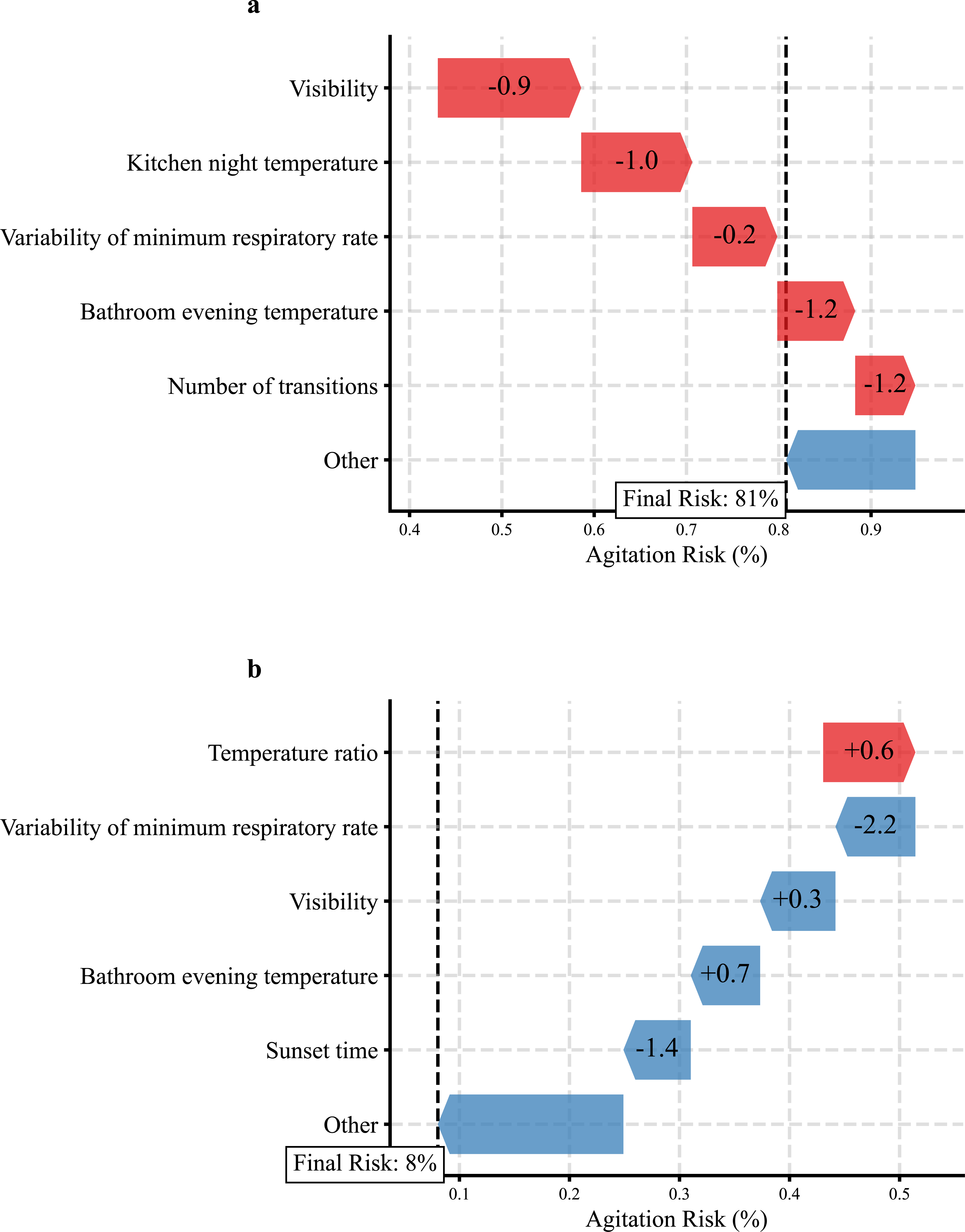
Personalised investigation of modifiable features. Examples from a positive (a) and a negative (b) event from PLwD A are shown using the SHAP framework. The colour of the arrow corresponds to the contribution: red contributes to agitation presence and blue contributes to agitation absence. Positive SHAP values contributed to positive predictions (agitated), while negative SHAP values contributed to negative predictions (non-agitated). The size of the arrow represents the absolute SHAP value, indicating the magnitude of each feature’s contribution. The number within the arrow corresponds to the normalised feature value.

To improve the clinical applicability of such analyses, we designed an interactive tool that allows clinicians to investigate the effect of contributing features at an individual level. The tool provides a visualisation of the predicted agitation risk and each feature’s contribution to the estimated risk (Figure 4).

**Figure 4:**
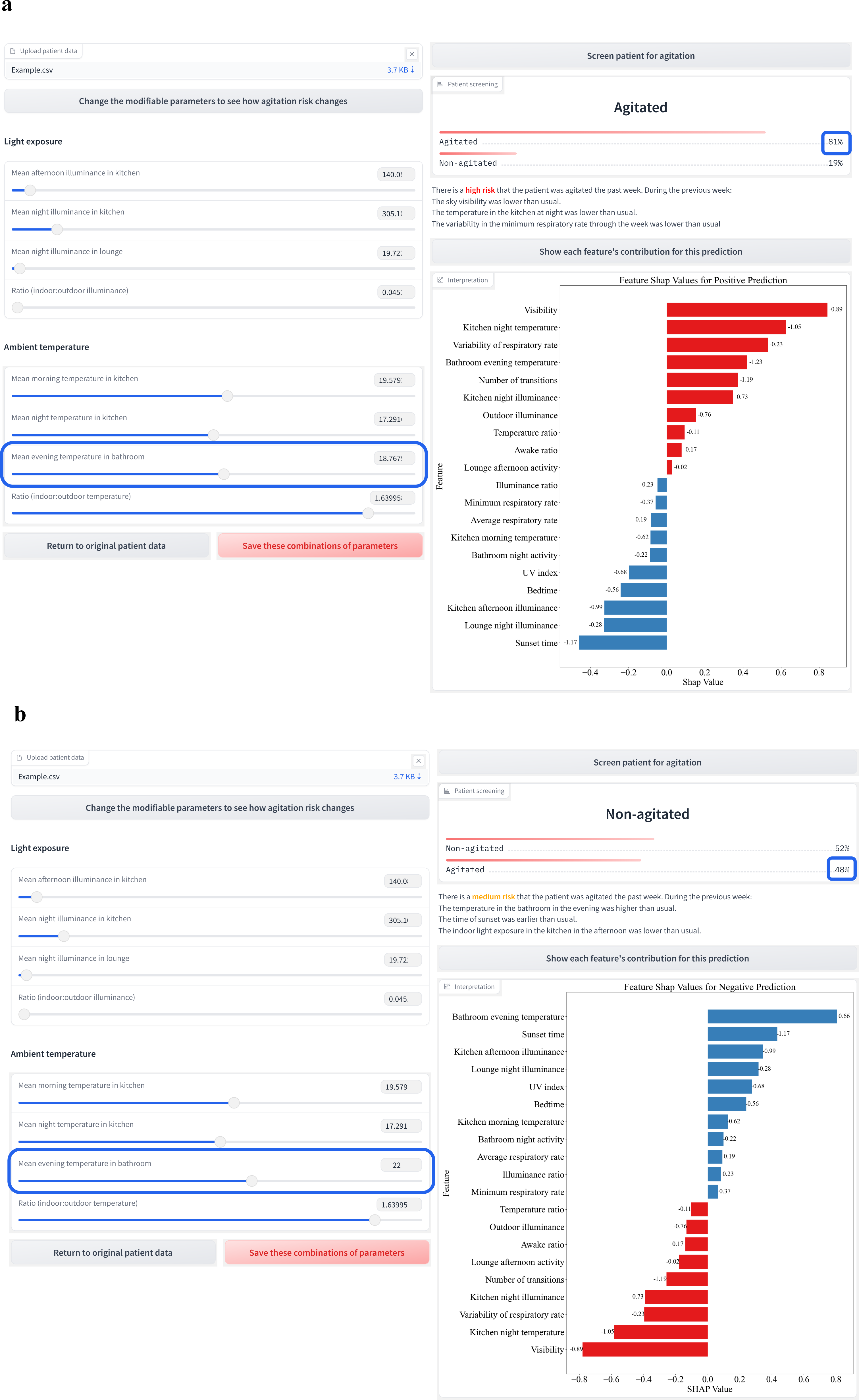
*In-silico* Experiment: Adjusting Temperature via an Interactive Interface. The interactive interface is shown, which accepts the input data as a CSV file. The tool provides sliding bars for the modifiable features and presents the predicted risk and associated probabilities, and a feature importance plot using SHAP values. In the feature importance plot, red bars correspond to features that contributed towards positive agitation prediction and blue bars correspond to features that predict the absence of agitation. Each bar is annotated with the corresponding normalised feature value. The user can save the combinations of modifications they have made to the modifiable parameters. a. Anonymised data from a participant. b. The results after modifying one of the parameters, morning indoor illuminance in the kitchen. An online version with a synthetic patient data generator is hosted on huggingface. (see https://huggingface.co/spaces/marirena/AgitationScreening). Access to the tool is currently restricted. Upon publication of our work, access will become open.

We used this interactive interface to perform *in-silico* experiments by exploring the effect of non-pharmacological interventions on the estimated risk of agitation. Through sliding bars, modifications of features can be simulated. The set of modifiable features investigated included 4 features on room temperature and 4 features on light exposure. By changing the values of these features, the model is re-run with the updated data, leading to a new agitation risk prediction. Exemplary, we demonstrated that by only modifying the evening indoor temperature in the bathroom from 18.77 to 22, the agitation risk for a participant who was initially identified as high risk (Red) changed from 81% to 48%, shifting them to medium risk (Amber) (Figure 4). Similarly, for a PLwD whose afternoon kitchen illuminance was identified as a risk factor (Figure S8), reducing the illuminance from 257.7 to 100 resulted in a risk decrease from 77% to 23% (Figure S9) shifting the risk from medium (Amber) to low (Green). Such *in-silico* experiments enable clinicians to investigate personalised interventions.

## Discussion

We used a unique dataset from an in-home monitoring dementia care study to identify and investigate the risk of agitation in PLwD. A LightGBM classifier was employed to ascertain weekly agitation risk. Integration of feature importance analysis alongside a traffic-light system significantly increased the clinical utility of this ML framework. The development of an interactive online platform facilitated a comprehensive *in-silico* examination of non-pharmacological interventions.

The retrospective analysis of weekly agitation risk, a novel approach within our study, provides opportunities to explore the underlying factors contributing to agitation. When comparing our model performance with existing literature, it is crucial to consider that our aim differed from that of previous models, which focused on detecting the onset of agitation. Khan *et al.*^11^ achieved a higher ROC-AUC score compared to our model (77.63% vs. 86.20%). However, their study had limited data available (n=2, labelled person-days=28) compared to ours (n=63, labelled person-days=4,096). Compared to our previous study by Palermo *et al.*^12^, our proposed model achieved lower sensitivity (71.36% vs. 79.78%) but higher precision and F1-score. This suggests a decrease in false alerts in our current model, increasing its clinical applicability. Our proposed model balanced both high sensitivity and specificity, ensuring sensitive screening, while reducing unnecessary alerts (Table 1, Figure S7).

Addressing fairness and bias, our analysis indicates an absence of bias towards specific groups within our cohort (see Table S8, Table S9, Figure S2), likely attributable to the personalised pre-processing employed. A large body of ML research currently focuses on developing accurate methods for personalised pre-processing^23^. Here, we scaled each feature to the baseline of each individual participant to accommodate underrepresented groups within our dataset. By considering individual baselines, we aimed to ensure that normal variations between groups were appropriately accounted for, without being conflated as indicators of agitation. This is the first study on ML applications for agitation that investigates model bias and model fairness. By doing so, we offer objective insights into this automated screening process.

Through rigorous feature analysis, our model’s interpretability and, by extension, its clinical utility have been significantly enhanced. Previous ML applications for agitation lacked insights into agitation-driving factors due to their more complex models. By incorporating a wide range of longitudinal data, including sleep, activity, and importantly, indoor and outdoor environmental factors, which have not been extensively investigated, our model revealed significant agitation indicators that are not captured in routine clinical practice. Agitation periods in our study exhibited a notably higher awake ratio during sleep, suggesting heightened alertness and decreased sleep quality. Additionally, we observed a significantly lower minimum nocturnal respiratory rate, potentially indicating nocturnal breathing cessations (Figure 2, Table S14). Extreme levels of light exposure, both high and low, were prevalent during agitation periods, alongside a significantly low illuminance ratio indicative of poor light quality (Figure 2, Table S21). Low temperatures in the bathroom were also present during agitation periods (Figure 2, Table S20). Our findings are consistent with previous clinical studies suggesting a link between sleep-disordered breathing and frequent awakenings with agitation^24,25^. Similarly, our observations align with prior research reporting associations of extremely low illuminance levels at night with falls and hallucinations in older adults, potentially exacerbating agitation symptoms^26,27^. Our finding of reduced light quality influencing agitation risk corresponds to an observational study reporting lower illuminance ratios in areas where PLwD tend to become agitated^28^. Our observation that significantly higher illuminance values contribute to agitation is supported by studies indicating that high illuminance can cause discomfort due to age-related eye sensitivity^29^. However, other studies that administered bright light therapy (BLT) to PLwD have shown mixed effects of high illuminance on agitation^30–32^. Notably, illuminance levels during agitation weeks in our study remained relatively dim, with the highest average being 455.18 lux (Table S15), contrasting with BLT protocols that often exceed 1000 lux^30–32^. Our study showed that lower indoor temperatures were correlated with agitation, which relates to a previous finding where a deviation from an optimal temperature of 22.6°C increased agitation behaviours^33^. Overall, our model identified several known sleep, illuminance and ambient temperature features associated with agitation, including nighttime disturbances^25^, sleep-disordered breathing^24^, poor light quality^28^ and low ambient temperature^33^. We further identified features that have not been previously associated with agitation in the literature, including extreme levels of outdoor visibility, pronounced differences between indoor and outdoor temperatures, as well as room and time-period specific illuminance and temperature measures, as illustrated in Figure 2.

Our interpretable ML framework enables the exploration of non-pharmacological strategies to mitigate agitation and improve the quality of life for PLwD. Such strategies include the diagnosis and treatment of coexisting sleep disorders, particularly disordered breathing. Additionally, minimising nighttime disturbances is vital for alleviating carer distress, thereby supporting patients to remain at home and avoid institutionalisation^34^. Our findings, along with current literature, suggest that modifiable parameters such as light exposure^35^ and temperature^33^ can be tailored for each PLwD to reduce the risk of agitation. Specifically, relying on natural light or adjusting artificial lighting to mimic natural light could improve light quality and alleviate agitation symptoms^35^. Furthermore, maintaining indoor temperatures within a moderate range^33^ and avoiding significant fluctuations could enhance comfort and help mitigate agitation episodes. Interventions targeting lighting levels in kitchen and lounge areas (Figure 4) and adjustments to bathroom and kitchen temperature could prove beneficial in our cohort (Figure 2).

This study highlighted ways to increase the clinical value of ML models through the inclusion of a traffic-light system and the development of an interactive interface, facilitating the exploration of personalised interventions. Identification of risk groups through a traffic-light system enables the effective management of urgent alerts (Red group), while minimising false alerts. The traffic-light system represents a comprehensive approach beyond binary predictions, a method not previously explored in other agitation screening models. This approach aligns with the NICE guidelines for monitoring serious diseases, highlighting its potential applicability in clinical practice^36^. The development of the first -to our knowledge- interactive tool for personalised intervention experimentation for agitation in dementia represents a significant advancement for dementia care. It aligns with current literature advocating for person-centred approaches to managing agitation in dementia^19^. By utilising this tool, clinicians can offer practical instructions to study partners for managing agitation.

The findings of this study are subject to limitations. Firstly, data quality may be compromised as participants reside with study partners (Table S1), potentially leading to the partners’ behaviour confounding the collected data. Future studies should use participant-specific data collection methods and investigate the role of study partners in assessing and managing agitation in PLwD. Secondly, the investigation of the potential effect of light exposure on behaviour is limited by only including illuminance data. Additional parameters such as spectral irradiance, bandwidth, and position of individuals relative to the light source have also been reported to influence behaviour^37^. Thirdly, due to the uncontrolled environment, other external parameters that are not captured by our data collection could be affecting the behaviour.

Despite the model’s fairness towards specific demographics within our cohort (Figure S2), it is crucial to acknowledge the lack of diversity in our dataset (Table S1). Addressing this limitation in future studies by recruiting participants from diverse demographic groups would not only ensure the fairness and generalisability of our framework across different populations but also allow for the external evaluation of our model.

Further limitations arise from the use of a weekly risk assessment approach. The labels derived from weekly reports might be inaccurate due to subjective and incomplete recollections of events. Nonetheless, our results demonstrate that employing ML models carefully and analysing longitudinal patterns allowed us to establish a reliable measure for identifying agitation patterns and episodes.

A further remark should be made on implied causality. Given our weekly approach to agitation detection, factors leading to agitation, alongside agitation behaviours, are included in the data. This could potentially render the identified markers epiphenomena rather than indicators of causality. To establish a causal relationship between environmental and sleep features, and agitation, controlled clinical intervention studies are needed. Our interface facilitates the identification of potential interventions to investigate.

Our next research phase involves conducting a clinical study to implement non-pharmaceutical interventions for agitation within our cohort, leveraging the developed model and tool. By employing this clinical decision-making support tool, we aim to assess its efficacy and clinical utility, while examining the effectiveness of the identified personalised interventions in managing agitation in dementia over the long term.

## Methods

### Study design and population

The Minder study was initiated in collaboration with Imperial College London, the University of Surrey and Surrey and Borders Partnership NHS Trust^20^. Eligible study participants included adults older than 50 years with a clinically ascertained diagnosis of dementia or mild cognitive impairment and current or previous treatment at a psychiatric unit. In total, 127 participants have been recruited (Table S1). Most participants live with carers or study partners who attend clinical assessments with them. More details on the study design are available in subsection S1.1.

### Data collection

Demographic data were collected during the baseline assessment. In-home monitoring data was continuously collected using low-cost sensors placed in the participants’ homes. These devices include passive infrared sensors (PIR), sleep mats, and door and kitchen appliance sensors. Here, we included data from the PIR sensors, comprising of activity, indoor light, and indoor temperature. We also integrated data from the sleep mat, which is placed underneath the mattress of each participant and monitors respiratory and heart rates, and nighttime events. Additionally, we included outdoor light, outdoor temperature, and weather data sourced from the Visual Crossing Weather API https://www.visualcrossing.com, specifically for the Surrey area where most enrolled participants resided. Lux illuminance units for outdoor light were derived from solar irradiance values, directly available on Visual Crossing, using the conversion factor of 122^38^. For a layout of all sensors used refer to Figure S1.

Clinical psychometric and cognitive assessment tools, including the Neuropsychiatric Inventory (NPI), were administered within the Minder study, to gather behavioural and cognitive data during regular (3-month) visits. Additionally, a weekly behavioural monitoring questionnaire process was implemented. During this weekly monitoring process, study partners and/or participants were contacted to report the presence or absence of participants’ behavioural symptoms regarding the preceding week. This included symptoms such as agitation, delusions, hallucinations, depression, and anxiety.

### Data labelling

In our study, agitation status was determined in a two-stage verification method, through the responses from the weekly monitoring process. Trained research staff, having completed both NIHR Good Clinical Practice (GCP) and Valid Informed Consent training, compiled unstructured notes based on their weekly interactions with carers/ participants and labelled the weeks for each participant as presence or absence of agitation. We conducted a review to confirm the correspondence of the notes with the labels (see Table S2). The whole preceding week, including the day of the reporting itself (8 days), was labelled as a binary target variable representing the presence or absence of agitation. An 8-day rolling window was applied to consistently extract 8 days of data preceding an event even if the time between events was smaller than 8. This approach was chosen to accurately simulate the model’s operation in real-world use. The weekly nature of the labels allowed for the observation of the period surrounding agitation episodes, facilitating the identification of broader agitation patterns and factors contributing to agitation. The weekly monitoring responses were chosen over the more detailed NPI ones, due to the weekly granularity enabling us to use the labels in association with passively collected sensor data. Using weekly labels further has the potential to reduce participant burden by minimising the frequency of contact for agitation events’ verification and questionnaire administration.

For our experimentation (Figure 1), we used the labelled data collected in the Minder study, from 15/07/2021 to 16/03/2023. To develop the ML model, we included individuals for whom the presence or absence of agitation episodes was recorded, leading to a final sub-cohort of 63 (41 male and 22 female participants) with a total of 242 positive and 270 negative agitation-labelled weeks. The demographics of the agitation sub-cohort are provided in Table S1.

### Data exploration and pre-processing

To determine which source of light and temperature (indoor and outdoor) data to include at different times during the day, we used PIR, door sensors, and the sleep mat to determine the location of the subject. Only indoor light and temperature values that corresponded to simultaneous (resolution = 1 hour) activity data from PIR sensors were retained. Outdoor values (daily averages) were retained only for days when there was an indication that the participant was not indoors. This was confirmed by only retaining days for which there was use of the front or back door and otherwise inactivity in the house. Indoor light, indoor temperature, and activity data were aggregated over time windows of 6 hours, representing different periods of the day: morning, afternoon, evening, and night, to capture fluctuations in environmental variables and activity signatures in the house throughout the day.

In our exploratory analysis, we conducted two-sided paired Student’s T-tests to compare various extracted measures describing sleep, illuminance and ambient temperature between agitation and non-agitation weeks. For this, we only included participants who had both positive and negative agitation events (n=29) (Table S1).

We handled each data modality separately, which involved retaining only those events for which data across all modalities were available. Missing indoor light, indoor temperature, sleep, and activity labelled data were imputed with the k-nearest neighbours algorithm (*k* = 5) separately for each participant. For missing values that remained, iterative imputation was performed using Bayesian Linear Ridge regression and initial strategy the median, with 10 iterations on unlabelled data from all participants. For the outdoor light and temperature data, when there was no outdoor activity suggested from the data due to challenges in integrating data from the different sources (activity, sleep, and door sensor), we used the average outdoor values from the entire week instead.

Z-score normalisation was performed to scale each participant’s data according to their individual baseline. This involved computing the individual baseline mean and standard deviation (SD), by considering both labelled and unlabelled data (32,896 person-days) collected from each participant throughout their entire participation in the Minder study. This scaling procedure was undertaken to mitigate potential individual differences among participants and ensure a standardised and fair comparison across the dataset. 7 participants in total lacked sufficient weekly data to compute reliable statistical properties for specific combinations of time-period and room features (e.g. Bathroom night illuminance or Kitchen night temperature). The missing data varied among participants, with each individual exhibiting gaps in different features. This variability stemmed from differences in home signature, behaviour, and duration of study participation among participants. To address this, we substituted the missing average or SD feature value with the average statistical properties derived from other participants.

### Feature Selection

We derived 114 variables from the sensor data, including sleep, activity, indoor and outdoor light exposure, indoor and outdoor temperature, and seasonality (Table S3). Feature selection was performed using the SHapley Additive exPlanation (SHAP)^22^ values, computed by a Random Forest Classifier (RF), using 10 folds. All data splits were stratified and were performed with a grouping strategy based on individual participant ID (ID-grouping), preventing data leakage. This resulted in 20 selected features considered for further modelling. More information on feature selection can be found in section S1.3.1.

### Machine learning models

To detect agitation, we compared the performance of RF^39^, Light Gradient-Boosting Machine (LightGBM)^40^ Extreme Gradient Boosting (XGBoost)^41^, Adaptive Boosting (ADABoost)^42^, Support Vector Machine (SVM)^43^, Logistic Regres-sion (LR)^44^ and a multilayer-perceptron (MLP). Details are provided in subsections S1.2, S1.3.

The hyperparameters for the models were tuned using grid search on stratified 5-Fold train/validation splits, with the model achieving the highest sensitivity on the validation data selected as the most suitable model (Details on hyperparameters are provided on Table S7). Model performance was evaluated using stratified, ID-grouping 10-Fold train/test splits. Sensitivity was prioritised as the primary metric to enhance the model’s capability to sensitively detect instances of agitation, as desired for medical applications. Performance of the 7 models on the 10 test sets, was compared using two-sided paired T-tests (N=10). The best-performing model (LightGBM) was identified and used for further analysis.

### Risk stratification

To further assist clinical decision-making, a traffic-light-based system was developed, which separated the agitation risk into three categories based on thresholds: Green (low risk), Amber (medium risk), and Red (high risk).

The thresholds were determined via a 10-fold nested stratified cross-validation approach on the LightGBM, incorporating ID-grouping to ensure that participants used to determine the thresholds were distinct from those used for evaluation. The inner 10-fold cross-validation determined thresholds based on validation set performance, while the outer 10-fold cross-validation evaluated model performance on independent test sets post-threshold application.

We optimised the traffic light model to have a balanced inclusion ratio between the Amber, Green and Red groups. By expanding the boundaries of the Amber group, the false alerts would be decreased and sensitivity would be increased. However, a disproportionate allocation of the results to the Amber group would decrease the specificity of the model.

To avoid this, we focused on jointly increasing sensitivity and specificity, by maximising Youden’s J index^45^. To achieve this, we varied the stratification thresholds with a resolution of 10% and computed the sensitivity and specificity of the predictions on the Green (no agitation predicted) and Red groups (agitation predicted) in the validation sets (Figure S5). To inform the range of the thresholds, we used a source different from the weekly labels used for model development to avoid circularity. Specifically, we used the NPI questionnaire responses, administered to the entire Minder cohort from July 2020 to March 2023, every three months (Table S10, subsection S1.4.3). The rate of Red (Rr) alerts was determined based on the agitation frequency domain reported in the NPI by the carers. The rate of Green alerts (Gr) was defined, due to agitation episodes being less frequent than their absence (see section S1.4.3). The thresholds were filtered based on the following criteria:

- Ensuring that the Rr is:

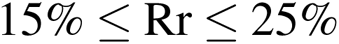

- Ensuring that the Gr is:

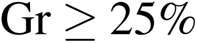

- Maximising Youden’s J index, defined as Sensitivity + Specificity — 1

When considering all thresholds adhering to these criteria on the validation sets, each average threshold was computed, resulting in 10 averages. During model evaluation on the test sets, the threshold from each respective validation set was used.

The average thresholds from all 10 folds were:

- Green: [0.00, 32.25]
- Amber: [32.25, 78.14]
- Red: [78.14, 100.00]

### Interactive interface

We created an interactive interface using the Gradio library^46^. The developed tool allows uploading patient data as a CSV file to compute weekly agitation risk based on the trained ML model. The corresponding risk group from the traffic-light system is displayed and feature contributions are shown using the SHAP values. We defined a set of modifiable factors: {Kitchen afternoon and night illuminance, Lounge night illuminance, illuminance ratio (average indoor:average outdoor), Kitchen morning temperature,Kitchen night temperature, Bathroom evening temperature, temperature ratio (average indoor:average outdoor}. These can be adjusted on the interface using sliding bars enabling a simulation of the effect of non-pharmacological interventions in a personalised framework. The user can download their applied modifications to facilitate the design of interventions. For our *in-silico* experimentation within this paper, we utilise the tuned and trained model from the corresponding fold to which the participant examined belongs in the test set. The tool is hosted on huggingface. The online version employs the LightGBM model tuned and trained on all data available from the 63 participants (https://huggingface.co/spaces/marirena/AgitationScreening). To ensure data privacy, we include a synthetic data generator in the online version. (Access to the tool is currently restricted. Upon publication of our work, access will become open.)

## Acknowledgements

This study is funded by the UK Dementia Research Institute [award number UK DRI-7002] through UK DRI Ltd, funded by the Medical Research Council (MRC), Alzheimer’s Research UK, Alzheimer’s Society, and the UKRI Engineering and Physical Sciences Research Council (EPSRC) PROTECT Project (grant number: EP/W031892/1). Infrastructure support for this research was provided by the NIHR Imperial Biomedical Research Centre (BRC) and the UKRI Medical Research Council (MRC).The funders were not involved in the study design, data collection, data analysis or writing the manuscript

## Data availability

Unidentified patient data can become available from the corresponding author upon reasonable request. The trained model is available at: https://huggingface.co/spaces/marirena/AgitationScreening/tree/main.

## Code availability

The code used in this study will be made available by the corresponding author upon reasonable request.

## Author Contributions

M.B.: Conceptualisation, Methodology, Software, Formal analysis, Investigation, Data Processing, Writing—Original Draft, Review and Editing, Visualisation; ANS, NFL: Methodology, Writing—Original Draft, Review and Editing; CW: Writing—Original Draft, Review and Editing, Data collection, AC: Provided visualisation package AVT, Reviewing and Editing CS, SK : Reviewing and Editing; RN: Clinical Study Lead, Conceptualisation, Data Collection, Writing—Review and Editing, Funding Acquisition; PB: Conceptualisation, Methodology, Writing—Original Draft, Review and Editing, Supervision, Funding Acquisition

## References

1. Sano, M. et al. Agitation in cognitive disorders: Progress in the International Psychogeriatric Association consensus clinical and research definition. International psychogeriatrics. 2023. 1–13. doi:10.1017/S1041610222001041.

2. Levy, M. L. et al. Longitudinal assessment of symptoms of depression, agitation, and psychosis in 181 patients with Alzheimer’s disease. The American journal of psychiatry. 1996. 153. 1438–1443. doi:10.1176/ajp.153.11.1438.

3. Cohen-Mansfield, J., Marx, M. S. & Rosenthal, A. S. A description of agitation in a nursing home. Journal of gerontology. 1989. 44. M77–84. doi:10.1093/geronj/44.3.m77.

4. Cohen-Mansfield, J. Measurement of inappropriate behavior associated with dementia. Journal of gerontological nursing. 1999. 25. 42–51. doi:10.3928/0098-9134-19990201-08.

5. Cerejeira, J., Lagarto, L. & Mukaetova-Ladinska, E. B. Behavioral and psychological symptoms of dementia. Frontiers in neurology. 2012. 3. 73. doi:10.3389/fneur.2012.00073.

6. Fillit, H. et al. Impact of agitation in long-term care residents with dementia in the United States. International journal of geriatric psychiatry. 2021. 36. 1959–1969. doi:10.1002/gps.5604.

7. Hiyoshi-Taniguchi, K., Becker, C. B. & Kinoshita, A. What Behavioral and Psychological Symptoms of Dementia Affect Caregiver Burnout? Clinical Gerontologist. 2018. 41. 249–254. doi:10.1080/07317115.2017.1398797.

8. Rosen, J. et al. The Pittsburgh Agitation Scale: A User-Friendly Instrument for Rating Agitation in Dementia Patients. The American journal of geriatric psychiatry : official journal of the American Association for Geriatric Psychiatry. 1994. 2. 52–59. doi:10.1097/00019442-199400210-00008.

9. Cohen-Mansfield, J. & Libin, A. Assessment of agitation in elderly patients with dementia: correlations between informant rating and direct observation. International Journal of Geriatric Psychiatry. 2004. 19. 881–891. doi:10.1002/gps.1171.

10. Finkel, S. I., Lyons, J. S. & Anderson, R. L. Reliability and validity of the Cohen–Mansfield agitation inventory in institutionalized elderly. International Journal of Geriatric Psychiatry. 1992. 7. 487–490. doi:10.1002/gps.930070706.

11. Khan, S. S. et al. *Agitation Detection in People Living with Dementia using Multimodal Sensors* in. 2019 41st Annual International Conference of the IEEE Engineering in Medicine & Biology Society (EMBC) (IEEE, Berlin, Germany, July 2019), 3588–3591. ISBN: 978-1-5386-1311-5. https://ieeexplore.ieee.org/document/8857781/.

12. Palermo, F. et al. Designing A Clinically Applicable Deep Recurrent Model to Identify Neuropsychiatric Symptoms in People Living with Dementia Using In-Home Monitoring Data in. NeurIPS 2021. Publisher: arXiv Version Number: 2 (2021). https://arxiv.org/abs/2110.09868.

13. HekmatiAthar, S., Goins, H., Samuel, R., Byfield, G. & Anwar, M. Data-Driven Forecasting of Agitation for Persons with Dementia: A Deep Learning-Based Approach. SN computer science. 2021. 2. 326. doi:10.1007/s42979-021-00708-3.

14. Al Ghassani, A., Rababa, M. & Abu Khait, A. Agitation in people with dementia: A concept analysis. Nursing Forum. 2021. 56. 1015–1023. doi:10.1111/nuf.12629.

15. Dementia: assessment, management and support for people living with dementia and their carers. (2018). NICE Guideline NG97.

16. Carrarini, C. et al. Agitation and Dementia: Prevention and Treatment Strategies in Acute and Chronic Conditions. Frontiers in neurology. 2021. 12. 644317. doi:10.3389/fneur.2021.644317.

17. Mitolo, M. et al. Effects of Light Treatment on Sleep, Cognition, Mood, and Behavior in Alzheimer’s Disease: A Systematic Review. Dementia and geriatric cognitive disorders. 2018. 46. 371–384. doi:10.1159/000494921.

18. Cheung, D. S. K. et al. Sensory-based interventions for the immediate de-escalation of agitation in people with dementia: A systematic review. Aging & Mental Health. 2023. 27. 1056–1067. doi:10.1080/13607863.2022.2116404.

19. James, I. A., Reichelt, K., Shirley, L. & Moniz-Cook, E. Management of Agitation in Behaviours That Challenge in Dementia Care: Multidisciplinary Perspectives on Non-Pharmacological Strategies. Clinical interventions in aging. 2023. 18. 219–230. doi:10.2147/CIA.S399697.

20. Ream, E. *MINDER Health Management Study for Dementia: A study to refine and evaluate technologies in the home to monitor and manage the health of people with dementia* Oct. 10, 2018. http://www.isrctn.com/ISRCTN71000991.

21. *Visual Crossing Weather* https://www.visualcrossing.com/ (2023).

22. Lundberg, S. & Lee, S.-I. A Unified Approach to Interpreting Model Predictions in Proceedings of the 31st International Conference on Neural Information Processing Systems (2017). arXiv: 1705.07874[cs, stat]. http://arxiv.org/abs/1705.07874.

23. Sharma, P., Shamout, F. E., Abrol, V. & Clifton, D. A. Data Pre-Processing Using Neural Processes for Modeling Personalized Vital-Sign Time-Series Data. IEEE journal of biomedical and health informatics. 2022. 26. 1528– 1537. doi:10.1109/JBHI.2021.3107518.

24. Karen M. Rose et al. Sleep Disturbances and Nocturnal Agitation Behaviors in Older Adults with Dementia. Sleep. 2011. doi:10.5665/SLEEP.1048.

25. Richards, K. C. et al. Nighttime Agitation in Persons with Dementia as a Manifestation of Restless Legs Syndrome. Journal of the American Medical Directors Association. 2021. 22. 1410–1414. doi:10.1016/j.jamda.2020.11.026.

26. Murgatroyd, C. & Prettyman, R. An investigation of visual hallucinosis and visual sensory status in dementia. International journal of geriatric psychiatry. 2001. 16. 709–713. doi:10.1002/gps.426.

27. Delepeleire, J., Bouwen, A., Deconinck, L. & Buntinx, F. Insufficient Lighting in Nursing Homes. Journal of the American Medical Directors Association. 2007. 8. 314–317. doi:10.1016/j.jamda.2007.01.003.

28. Kenji, N. & Neveen, H. *Assessment Of Daylight In Relation To The Agitation Levels Of People With Dementia* in *BSO Conference* Proceedings of BSO Conference 2016: Third Conference of IBPSA-England. **3**. Journal Abbreviation: BSO Conference (IBPSA-England, Newcastle, UK, Sept. 2016), 170–177. https://publications.ibpsa.org/conference/paper/?id=bso2016_1013.

29. Konis, K., Mack, W. J. & Schneider, E. L. Pilot study to examine the effects of indoor daylight exposure on depression and other neuropsychiatric symptoms in people living with dementia in long-term care communities. Clinical interventions in aging. 2018. 13. 1071–1077. doi:10.2147/CIA.S165224.

30. Barrick, A. L. et al. Impact of ambient bright light on agitation in dementia. International journal of geriatric psychiatry. 2010. 25. 1013–1021. doi:10.1002/gps.2453.

31. Schindler, S. D., Graf, A., Fischer, P., Tolk, A. & Kasper, S. Paranoid delusions and hallucinations and bright light therapy in Alzheimer’s disease. International journal of geriatric psychiatry. 2002. 17. 1071–1072. doi:10.1002/gps.497.

32. Burns, A., Allen, H., Tomenson, B., Duignan, D. & Byrne, J. Bright light therapy for agitation in dementia: a randomized controlled trial. International psychogeriatrics. 2009. 21. 711–721. doi:10.1017/S1041610209008886.

33. Tartarini, F., Cooper, P., Fleming, R. & Batterham, M. Indoor Air Temperature and Agitation of Nursing Home Residents With Dementia. American journal of Alzheimer’s disease and other dementias. 2017. 32. 272–281. doi:10.1177/1533317517704898.

34. Vitiello, M. V. & Borson, S. Sleep Disturbances in Patients with Alzheimer??s Disease: Epidemiology, Pathophysiology and Treatment. CNS Drugs. 2001. 15. 777–796. doi:10.2165/00023210-200115100-00004.

35. Saidane, H. A., Rasmussen, T., Andersen, K., Iversen, H. K. & West, A. S. An Explorative Investigation of the Effect of Naturalistic Light on Agitation-Associated Behavior in Nursing Home Residents With Dementia: A Pilot Study. HERD. 2023. 16. 146–154. doi:10.1177/19375867221146154.

36. Fever in under 5s (2014) NICE Quality standard 64. Last updated 27 July 2022.

37. Spitschan, M. et al. How to Report Light Exposure in Human Chronobiology and Sleep Research Experiments. Clocks & Sleep. 2019. 1. 280–289. doi:10.3390/clockssleep1030024.

38. Michael, P. R., Johnston, D. E. & Moreno, W. A conversion guide: solar irradiance and lux illuminance. Journal of Measurements in Engineering. 2020. 8. 153–166. doi:10.21595/jme.2020.21667.

39. Breiman, L. Random Forests. Machine Learning. 2001. 45. 5–32. doi:10.1023/A:1010933404324.

40. Ke, G. et al. LightGBM: A Highly Efficient Gradient Boosting Decision Tree in. Advances in Neural Information Processing Systems (eds Guyon, I. et al.) **30** (Curran Associates, Inc., Long Beach, CA, USA, 2017), 3146–3154. https://proceedings.neurips.cc/paper_files/paper/2017/file/6449f44a102fde848669bdd9eb6b76fa-Paper.pdf.

41. Chen, T. & Guestrin, C. *XGBoost: A Scalable Tree Boosting System* in *Proceedings of the 22nd ACM SIGKDD International Conference on Knowledge Discovery and Data Mining* KDD ’16: The 22nd ACM SIGKDD International Conference on Knowledge Discovery and Data Mining (ACM, San Francisco California USA, Aug. 13, 2016), 785–794. ISBN: 978-1-4503-4232-2. https://dl.acm.org/doi/10.1145/2939672.2939785.

42. Schapire, R. E. in *Empirical Inference* 37–52 (Springer, Berlin, Heidelberg, 2013). ISBN: 978-3-642-41135-9 978-3-642-41136-6. https://link.springer.com/10.1007/978-3-642-41136-6_5.

43. Cortes, C. & Vapnik, V. Support-vector networks. Machine Learning. 1995. 20. 273–297. doi:10.1007/BF00994018.

44. The Regression Analysis of Binary Sequences. 20.

45. Youden, W. J. Index for rating diagnostic tests. Cancer. 1950. 3. 32–35. doi:10.1002/1097-0142(1950)3:1<32::AID-CNCR2820030106>3.0.CO;2-3.

46. Abid, A. et al. Gradio: Hassle-Free Sharing and Testing of ML Models in the Wild in ICML : Workshop on Human In the Loop Learning (HILL) Publisher: arXiv Version Number: 1 (Long Beach, California, USA, 2019). https://arxiv.org/abs/1906.02569.

