## Supplemental Appendix for "An Interpretable Machine Learning Tool for In-Home Screening of Agitation Episodes in People Living with Dementia"

### Table of Contents


S1.1 Study design and data sources 3
Table S1: Demographics 3
Figure S1: Data sources layout 3
Table S2: Example notes and labelling from the weekly monitoring process 3
S1.2 Development tools 3
S1.3 Model selection and optimisation 3
S1.3.1 Features and feature selection 4
Table S3: Features 4
Table S4: Performance Metrics of Feature Selection Models 5
Table S5: Performance Metrics Of Agitation Detection Models 5
Table S6: Statistical Results for the Comparison of Performance Metrics between LightGBM and baseline models 5
Table S7: Grid Search Hyperparameters 5
S1.4 Model evaluation 5
S1.4.1 Assessing Model Fairness and Bias 5
Table S8: Comparison of Model Performance between Males and Females 5
Table S9: Demographics of Consistently Correct and Incorrect Predictions 5
Figure S2: Bias analysis 5
S1.4.2 Reliability & Calibration 7
Figure S3: Calibration plot 7
Figure S4: Reliability plot 7
S1.4.3 Risk stratification 7
Table S10: Estimation of Agitation Frequency 7
Figure S5: Determination of Risk Stratification Thresholds 7
Figure S6: Effect of Risk Stratification 7
Figure S7: Precision-Recall (PR) and Receiver-Operating Characteristic (ROC) Curves for the Gradient Boosting (LightGBM) binary classification and classification after traffic-light based stratification 7
S1.5 Performance with fewer sensors 8
Table S11: Performance of Light Gradient Boosting (LightGBM) Classifier on Feature Subsets 8
S1.6 Explainability 8
Figure S8: Personalised investigation of modifiable features using SHAP local explanations. 8
Figure S9: *In-silico* Experiment: Adjusting Light Exposure via an Interactive Interface 8
S1.7 Supporting statistical analysis 8
Table S12: Measures used in statistical analysis 9
S1.7.1 Poor nighttime sleep and irregular sleep patterns as agitation indicators 9
Figure S10: Sleep measures comparison 9
Figure S11: Sleep measures variability comparison 9
Table S13: Descriptive Statistics for Sleep Measures 10
Table S14: Statistical results for the Comparison of Sleep Measures between agitation and non-agitation weeks 10
S1.7.2 Increased indoor lighting and poor light quality as indicators of increased agitation risk 19
Figure S12: Light Exposure Measures Comparison 19
Figure S13: Comparison of Combination of Indoor and Outdoor Light Exposure Measures 19
Table S15: Descriptive Statistics for the Comparison of indoor Illuminance Measures for agitation and non-agitation weeks. 19
Table S16: Statistical Results for the Comparison of Indoor Illuminance Measures 19
Table S17: Descriptive Statistics for the Comparison of Outdoor Illuminance Related Measures 26
Table S18: Statistical Results for the Comparison of indoor Illuminance Measures 26
Table S19:: Descriptive Statistics for the Comparison of indoor Temperature Measures 26

Table S20: Statistical Results for the Comparison of Indoor Temperature Measures 26
Table S21: Statistical Results for the Comparison of Outdoor Ambient Related Measures 26

#### S1.1 Study design and data sources

The study received ethical approval from the London-Surrey Borders Research Ethics Committee; TIHM 1.5 REC 19/LO/0102. The study is registered with the National Institute for Health and Care Research (NIHR) in the United Kingdom under the Integrated Research Application System (IRAS) registration number 257561.

Participants in the Minder study were recruited from: (1) health and social care partners within the primary care network and community NHS trusts, (2) urgent and acute care services within the NHS, (3) social services who oversee sheltered and extra care sheltered housing schemes, (4) NHS Community Mental Health Teams for older adults (CMHT-OP), and (5) specialist memory services at Surrey and Borders Partnership NHS Foundation Trust.

Participants lacking capacity for informed consent were required to have a study partner or carer who had known them for at least 6 months and was able to attend clinical study assessments with them. Exclusion criteria were as follows: (1) patients receiving treatment for terminal illness (2) presence of severe mental health conditions (3) presence of active suicidal thoughts.

Participants received an information sheet outlining how their personal data would be used in compliance with GDPR requirements. All participants provided written informed consent. The capacity to consent was assessed according to Good Clinical Practice, as detailed in the Research Governance Framework for Health and Social Care (Department of Health 2005) and the Mental Capacity Act 2005. If a participant lacking capacity expressed willingness to participate, a personal consultee would sign a declaration of consent. If no personal consultee was available, a professional consultee such as a key worker was sought. This process was detailed in the study protocol and approved by the ethics panel[1].

Capacity of both the participant and study partner is assessed at each research visit. The research staff conducting the assessment have completed the NIHR Good Clinical Practice (GCP) training and the Valid Informed Consent training. The demographics of the participants recruited in the Minder study are shown in Table S1.

The Minder study is an in-home monitoring study which employs low-cost in-home passive sensors, including PIR sensors, sleep mats and door sensors. The layout of the in-home monitoring devices employed in the Minder study[1], alongside an external sensor data source — the Visual Crossing Weather API (`https://www.visualcrossing.com`) that were used in the present study is shown in Figure S1.

In our study, agitation status was determined through

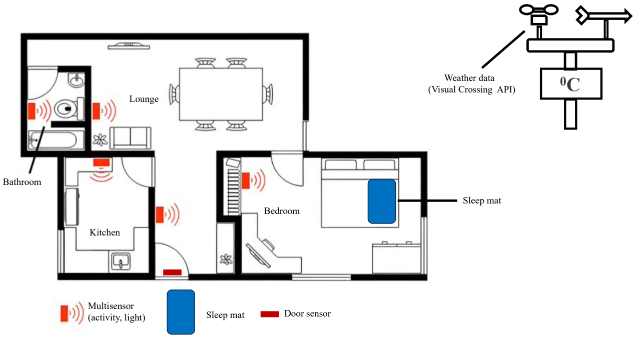


**Figure S1: Data sources layout.** Left: in-home monitoring devices (Minder study). Right: outdoor data from Visual Crossing Weather API (`https://www.visualcrossing.com`).

the responses from the weekly monitoring process, conducted by trained research staff, having completed both NIHR GCP and Valid Informed Consent training. Examples from the notes and the labelling can be seen in Table S2.

#### S1.2 Development tools

All pre-processing steps and modelling were performed using Python (3.9.12). The libraries Pandas (1.4.0)[2], NumPy (1.23.0)[3], SciPy (1.13.0)[4], Scikit-learn (1.4.0)[5] and TensorFlow(2.12.0)[6] were used for analysis and ML modelling. The Seaborn (0.11.2)[7] and Matplotlib (3.5.0)[8] libraries were used for visualisations. The SHAP (0.44.0) library was used for SHAP value computation and visualisations. The statistical testing was performed using the package pingouin (0.5.3)[9]. The interactive interface was created using the open-source package Gradio (4.21.0)[10].

#### S1.3 Model selection and optimisation

All models were evaluated with a stratified train-test split to ensure a similar proportion of agitation events across splits. Data leakage was prevented by ensuring that participants with multiple labels only occurred in either the training or the test set (ID-grouping). We reported several metrics for model performance: sensitivity, specificity, precision, area under precision-recall curve (PR AUC), area under receiver operator curve (ROC AUC), accuracy, and F1 score.

During model training, we prioritised maximising sensitivity to optimise our screening tool's effectiveness for sensitive agitation detection, as required in medical applications. When optimising the final model via risk strati-

**Table S1: Demographics.** Demographics of participants in the entire minder cohort (n=127), participants that were used in the agitation analysis (n=63) and participants used in the statistical analysis (n=29) are shown. The number of participants in each group is displayed.

| Characteristic | Entire Minder Cohort | Agitation Cohort | Statistical Analysis Cohort |
|---|---|---|---|
| **Total** | 127 | 63 | 29 |
| **Diagnosis** | | | |
| Alzheimer's disease | 59 | 36 | 14 |
| Vascular dementia | 7 | 5 | 3 |
| Frontotemporal dementia | 6 | 1 | 1 |
| Parkinson's disease | 4 | 3 | 2 |
| Unspecified dementia | 31 | 13 | 7 |
| Other and mixed | 20 | 5 | 2 |
| **Age Group** | | | |
| 50-60 | 1 | 0 | 0 |
| 60-70 | 12 | 7 | 1 |
| 70-80 | 29 | 13 | 7 |
| 80-90 | 55 | 31 | 18 |
| 90-100 | 27 | 12 | 3 |
| N/A | 3 | 0 | 0 |
| **Gender** | | | |
| Male | 69 | 41 | 23 |
| Female | 57 | 22 | 6 |
| Unspecified | 1 | 0 | 0 |
| **Ethnicity** | | | |
| White | 100 | 52 | 23 |
| Asian/Asian British | 8 | 4 | 2 |
| Black/African/Caribbean/Black | 3 | 1 | 0 |
| Mixed/Multiple ethnic groups | 1 | 0 | 0 |
| N/A | 15 | 4 | 4 |
| **Household** | | | |
| Multiple occupancy | 71 | 49 | 25 |
| Single occupancy | 49 | 13 | 3 |
| N/A | 7 | 1 | 1 |

fication, we aimed for a balance between sensitivity and specificity to avoid false alerts. The combination of the two metrics as Youden's J statistic was used for the risk stratification. It is calculated as:

$$J = \text{Sensitivity} + \text{Specificity} - 1. \quad (1)$$

This statistic ranges from 0 to 1, where a higher value indicates a better overall performance.

###### S1.3.1 Features and feature selection

The initial dataset included 114 features described in Table S3.

To prevent overfitting and improve model generalisability, feature selection was performed. We used an embedded method, as it is less computationally expensive than wrapper selection methods[11] and offers accuracy and interpretability when combined with the SHAP framework[12]. Further, the selected method was preferred over filter feature selection methods, since those only examine the univariate relationship between agitation and each feature, which does not reflect real-world scenarios, characterised by interaction between variables. In our embedded method, we combined ensemble tree-based models and the SHAP framework. We explored RF, XGBoost, ADABoost, and LightGBM classifiers. Their hyperparameters were tuned with grid search (Table S7) on 5 stratified cross-validation splits, with sensitivity as the scoring metric. The performance of these models was compared using stratified, ID-grouping 10-Fold cross-validation on the dataset contain-

**Table S2: Example notes and labelling from the weekly monitoring process.** Agitation status was determined through responses from the study partners or PLwD during the weekly monitoring process. The monitoring team documented notes from the monitoring process and labelled each week for every participant (negative: no agitation, positive: agitation) after reviewing those notes. Words within square brackets have been added to the original notes for better understanding. Notes with ... before or after indicate that some information has been omitted for privacy purposes. The keywords identified during labelling are highlighted in bold.

| Label | Monitoring process notes |
|---|---|
| Positive | - PLwD is getting very **irritable** and **agitated** that they cannot hear people. Carer believes this is because they cannot concentrate during conversations or get confused as opposed to hearing. Carer reported that PLwD is being very **rude** to formal carers...<br>- PLwD has become **irritable/ agitated** a bit more frequently recently. Some [symptoms] are not new, like reminding PLwD that they cannot drive anymore. Carer feels that PLwD requires lots of attention though, especially conversing with others. Otherwise PLwD can feel **irritated**.<br>- PLwD **irritated** with carer - professional. PLwD insists they can wash themselves but carer says they do need help.PLwD paranoid about who comes into the house. **Irritated** with inconsistencies [in conversations with carer]...<br>- Carer says PLwD has been very **irritable** and irrational in the last few days, they don't know of anything that has particularly set [the PLwD] off.<br>- Symptoms ongoing, **agitated**, trying...to get out of the house. |
| Negative | - Carer reports **no agitation**, irritability, or aggression on their part, and they are not distressed by the situation. |

ing all features (Table S3). RF performed best and was thus applied for the final feature selection.

SHAP values were computed using RF. Features with SHAP values greater than 0.015347 were retained, resulting in the selection of the top 20 features (The 21st feature had a SHAP value of 0.014600). The number was selected to increase interpretability while ensuring good predictive performance. The selected 20 features were used for subsequent training of models for agitation risk prediction.

One of the examined ML models was an MLP. This had the following architecture: number of input features, 32, 16, 8, 1. We chose the exponential Linear Unit (eLU) as the activation function, as it can handle negative input. The last layer employed sigmoid activation. The batch size was 32, and the maximum number of epochs was 50.

#### S1.4 Model evaluation

##### S1.4.1 Assessing Model Fairness and Bias

We investigated potential biases of the model by dividing the results by gender (number of females = 22). The model achieved lower performance for female participants, with decreased precision (7%) and specificity (2%) (Table S8). This indicates that the model incorrectly flagged females as agitated more frequently in comparison to males.

We also investigated biases for different dementia diagnoses, living conditions, and age. This was essential given the varied composition of our cohort (Table S1). Figure S2 illustrates a consistent ratio of prediction outcomes -True Positives (TP), True Negatives (TN), False Positives (FP), False Negatives (FN)- in various demographics. The absence of certain prediction outcomes within specific demographic groups, such as the 'No data' group in Figure S2.c and the 'Frontotemporal dementia' group in Figure S2.d, could be attributed to the limited number of events associated with participants in these particular groups.

Identifying participants for whom the model consistently provided accurate or inaccurate predictions was crucial to assess potential biases towards specific groups. Table S9 reveals that there was no specific pattern among participants for whom the model predicted inaccurately.

**Table S3: Features.** For each category, the number of features considered for feature selection alongside examples are listed.

| Feature Category | Number of features | Examples |
|---|---|---|
| Sleep | 25 | Ratio of sleep spent in each sleep state (awake, light, REM, deep), physiological variables while asleep (minimum, average, maximum heart rate, respiratory rate), bedtimes, waketimes |
| Sleep Variables Fluctuation (SD) | 25 | Standard deviation (SD) of the above sleep variables |
| Activity | 17 | Transitions, room-specific and time-specific activity (minutes spent) (e.g., kitchen morning activity) |
| Light Exposure | 20 | Room-specific and time-specific light exposure (e.g. kitchen morning temperature), outdoor illuminance, UV index |
| Temperature | 24 | Room-specific and time-specific (e.g., kitchen morning temperature) indoor temperature, outdoor temperature (average, minimum, maximum), apparent outdoor temperature (average, minimum, maximum) |
| Seasonality | 3 | Visibility, Cloud cover, Sunset time |
| **Total** | **114** | |

**Table S4: Performance Metrics of Feature Selection Models.** Comparison of tree-based models for feature selection on stratified, ID-grouping 10-Fold cross-validation. Performance metrics are reported as mean ± standard deviation. Models include ADABoost (Adaptive Gradient Boosting), LightGBM (Light Gradient Boosting Machine), RF (Random Forest), and XGBoost (Extreme Gradient Boosting). Metrics include ROC AUC (area under the receiver operating characteristic curve), PR AUC (area under the precision-recall curve), sensitivity, precision, specificity, accuracy, and F1 score. The best-performing model per metric is highlighted in bold.

| Model | Accuracy | F1 Score | Precision | Sensitivity | Specificity | PR AUC | ROC AUC |
|---|---|---|---|---|---|---|---|
| **ADABoost** | $56.38 \pm 8.93$ | $55.61 \pm 8.80$ | $57.27 \pm 9.81$ | $56.38 \pm 9.12$ | $61.01 \pm 16.61$ | $62.66 \pm 5.87$ | $58.19 \pm 7.61$ |
| **XGBoost** | $47.10 \pm 7.70$ | $40.25 \pm 7.36$ | $50.07 \pm 16.23$ | $46.84 \pm 6.18$ | $57.6 \pm 34.19$ | $45.79 \pm 7.70$ | $44.28 \pm 9.70$ |
| **RF** | $\mathbf{61.95 \pm 8.68}$ | $\mathbf{60.65 \pm 8.69}$ | $\mathbf{63.54 \pm 10.67}$ | $\mathbf{61.54 \pm 8.92}$ | $\mathbf{71.31 \pm 15.93}$ | $\mathbf{68.62 \pm 8.79}$ | $\mathbf{64.9 \pm 11.21}$ |
| **LightGBM** | $60.04 \pm 8.23$ | $59.47 \pm 8.59$ | $60.38 \pm 8.46$ | $60.09 \pm 8.37$ | $61.57 \pm 12.87$ | $69.63 \pm 9.2$ | $66.98 \pm 9.42$ |

**Table S5: Performance Metrics Of Agitation Detection Models.** Comparison of classifier performance on stratified, ID-grouping, 10-Fold cross-validation, using the 20 selected features. The performance metrics are reported as mean ± standard deviation. The different models compared are ADABoost (Adaptive Gradient Boosting), LightGBM (Light Gradient Boosting Machine), RF (Random Forest), XGBoost (Extreme Gradient Boosting), SVM (Support Vector Machine), LR (Logistic Regression), and an MLP (Multi-Layer Perceptron). Metrics include ROC AUC (area under the receiver operating characteristic curve), PR AUC (area under the precision-recall curve), sensitivity, precision, specificity, accuracy, and F1-score. The best-performing model is highlighted in bold.

| Model | Accuracy | F1 Score | Precision | Sensitivity | Specificity | PR AUC | ROC AUC |
|---|---|---|---|---|---|---|---|
| **ADABoost** | $62.94 \pm 5.89$ | $62.26 \pm 5.90$ | $63.41 \pm 7.05$ | $62.61 \pm 6.07$ | $70.59 \pm 10.91$ | $68.68 \pm 6.68$ | $67.47 \pm 6.65$ |
| **XGBoost** | $56.19 \pm 5.95$ | $45.09 \pm 14.14$ | $70.88 \pm 8.16$ | $54.72 \pm 6.90$ | $78.19 \pm 32.21$ | $57.23 \pm 12.32$ | $60.36 \pm 10.43$ |
| **RF** | $64.88 \pm 5.92$ | $63.96 \pm 5.73$ | $66.30 \pm 7.59$ | $64.54 \pm 5.89$ | $73.26 \pm 13.10$ | $74.84 \pm 7.71$ | $73.99 \pm 8.35$ |
| **LightGBM** | $\mathbf{71.36 \pm 7.21}$ | $\mathbf{71.12 \pm 7.28}$ | $\mathbf{71.81 \pm 7.94}$ | $\mathbf{71.32 \pm 7.38}$ | $\mathbf{75.28 \pm 10.43}$ | $\mathbf{78.6 \pm 7.57}$ | $\mathbf{77.63 \pm 6.59}$ |
| **LR** | $58.67 \pm 8.48$ | $57.4 \pm 8.4$ | $59.21 \pm 9.93$ | $58.01 \pm 8.28$ | $67.9 \pm 13.97$ | $64.49 \pm 8.57$ | $64.32 \pm 9.37$ |
| **SVM** | $64.91 \pm 7.78$ | $64.51 \pm 7.69$ | $65.23 \pm 7.93$ | $64.75 \pm 7.63$ | $67.02 \pm 12.87$ | $68.85 \pm 8.5$ | $69.39 \pm 9.5$ |

##### S1.4.2 Reliability & Calibration

We investigated the reliability and calibration of our models. The reliability plot shows the confidence surrounding the model's predictions (Figure S4). The model was reliable when predicting both positive and negative episodes of agitation. The ratio of predictions around extreme probabilities, 0.0 and 1.0, was low (Figure S3) which can be attributed to LightGBM classifier being an ensemble method, where the outcome is a combination of several decision trees. The model's calibration was overall good, with the average calibration curve closely aligning with that of a perfect classifier.

##### S1.4.3 Risk stratification

We incorporated risk stratification to increase the clinical value of our model. We achieved this by categorising the agitation predictions into three distinct risk groups based on a traffic-light system. We defined three risk groups (Red = high risk, Amber = moderate risk, Green = low risk) based on the Red and Green thresholds. The Amber group was then defined by the range between the Red and Green thresholds.

To inform the ranges of these thresholds, we investigated the prevalence of weekly agitation episodes based on the NPI scores. We analysed data from the whole Minder cohort, which included a total of 508 NPI questionnaires administered from July 2020 to March 2023. On average, each participant completed 4.10 ± 2.45 questionnaires. Of these, 118 questionnaires were from the baseline/first assessment. We report the frequencies of agitation symptoms in the baseline and all subsequent assessments in Table S10.

We grouped participants with no reported agitation and those who had reported agitation less than once per week. The remaining participants were aggregated. Subsequently, we calculated the ratio of the groups to receive an estimate of the prevalence of agitation. In the baseline assessment, the prevalence of agitation occurring at least once per week was 15%, while in the total of the follow-up assessments it was 23%. Taking into account the prevalence observed in our dataset (Table S10), we set the prevalence of Red alerts to range from 15% to 25% when defining the risk stratification thresholds.

The process of determining the thresholds for the three risk groups (Red = high risk, Amber = moderate risk, Green = low risk) involved exploring various combinations of Red and Green thresholds in the validation sets. Figure S5 illustrates how metrics and the percentage of predictions in the Red and Green groups fluctuated with varying thresholds applied to the validation sets. This shows how the optimal threshold was chosen using the nested K-fold. All the threshold combinations of the validation sets were filtered based on the Rr and Gr and Youden's J index (filtering out all rows (combinations of thresholds) that achieve Youden's J < 0.4).

After the application of the thresholds in the test sets, all metrics improved (see Table 1). The PR-AUC and ROC curves further show an improvement in the model (Figure S7).

**Table S6: Statistical Results for the Comparison of Performance Metrics between Light Gradient Boosting Machine classifier (LightGBM) and baseline models** Results of paired T-tests for comparison of sensitivity and specificity across 10 Folds between LightGBM and the following models: Adaptive Boosting (ADABoost), Extreme Gradient Boosting (XGBoost), Random Forest (RF), Logistic Regression (LR), Support Vector Machine (SVM), and a Multilayer Perceptron (MLP). The uncorrected p-values are reported. Significant results ($p - value < 0.05$) are highlighted in bold.

| Model | Metric | T-statistic | p-value | CI95% | Cohen-D | dof | Power |
|---|---|---|---|---|---|---|---|
| **RF** | Recall | 3.62 | 0.006 | [0.03, 0.11] | 0.99 | 1,9 | 0.79 |
| | Specificity | 0.62 | 0.55 | [-0.05, 0.09] | 0.17 | 1,9 | 0.08 |
| **ADABoost** | Recall | 4.44 | 0.002 | [0.04, 0.13] | 1.25 | 1,9 | 0.94 |
| | Specificity | 2.17 | 0.06 | [-0.0, 0.10] | 0.43 | 1,9 | 0.23 |
| **XGBoost** | Recall | 7.92 | 0.000 | [0.12, 0.21] | 2.29 | 1,9 | 1.00 |
| | Specificity | -0.25 | 0.81 | [-0.29, 0.23] | 0.12 | 1,9 | 0.06 |
| **LR** | Recall | 4.78 | 0.000 | [0.07, 0.20] | 1.68 | 1,9 | 1.00 |
| | Specificity | 1.83 | 0.10 | [-0.02, 0.17] | 0.59 | 1,9 | 0.39 |
| **SVM** | Recall | 2.08 | 0.07 | [-0.01, 0.14] | 0.86 | 1,9 | 0.68 |
| | Specificity | 1.92 | 0.09 | [-0.01, 0.18] | 0.70 | 1,9 | 0.50 |
| **MLP** | Recall | 2.03 | 0.07 | [-0.01, 0.12] | 0.80 | 1,9 | 0.62 |
| | Specificity | 2.33 | 0.04 | [0.00, 0.21] | 0.80 | 1,9 | 0.61 |

#### S1.5 Performance with fewer sensors

To address computational costs and energy consumption, an investigation was conducted to determine whether certain sensors could be omitted from the model. The process involved selecting the 20 most important features from each of the seven different sensor-specific subsets of the original dataset. The activity dataset only had 17 features available, so feature selection was omitted here (Table S3). A LightGBM classifier was trained with stratified 10-Fold train/test split using the seven different subsets of features (Table S11). The highest sensitivity and specificity scores were achieved when features from all sensors were integrated (Table S11). The lowest performance was observed when only activity features were considered. Notably, the model only showed a minimal decline in performance when activity was omitted.

#### S1.6 Explainability

To enhance the clinical utility of our model, we leveraged the local explanations provided by the SHAP framework to identify individual agitation risk factors for each PLwD. Additionally, we developed an interactive interface, offering a user-friendly version of the ML screening model, to facilitate the investigation of personalized interventions.

Using the SHAP framework, we identified afternoon kitchen illuminance as an agitation contributor for a PLwD. Figure S8 By reducing the illuminance from 257.7 to 100 there was a decrease in risk from 77% to 23% Figure S9. This change shifted the PLwD agitation risk from medium (Amber) to low (Green).

#### S1.7 Supporting statistical analysis

During our exploratory analysis, we derived weekly measures for sleep quality, sleep fluctuation, and indoor, and outdoor light exposure and ambient temperature, and compared them between agitation and non-agitation weeks. The measures derived are summarised in Table S12.

We performed paired T-tests to compare the same participant during agitation and non-agitation events. We thus, only retained participants with data available for both agitation and non-agitation weeks. The number of participants for each analysis will be shown using degrees of freedom (DDOF) when presenting the statistical results. The p-values reported are uncorrected.

The weekly average of all daily means was computed. For sleep measures, measures describing sleep variability, time-specific / room-specific indoor illuminance and temperature (e.g., illuminance in the kitchen at night), and the ratio of indoor to outdoor illuminance and temperature, z-score normalisation was performed per participant across positive and negative events. This approach aimed to account for individual differences and minimise bias towards agitation episodes. Outdoor illuminance and temperature were z-score normalised across all participants (due to no individual differences) and across both positive and neg-

**Table S7: Grid Search Hyperparameters.** Hyperparameter values used in Grid Search for hyperparameter tuning for different models. All hyperparameters not explicitly mentioned are set to their default values provided by scikit-learn (1.4.1)

| Hyperparameter | RF | XGBoost | LightGBM | ADABoost | MLP | SVM | LR |
|---|---|---|---|---|---|---|---|
| Number of Estimators | [50, 200, 500] | [50, 100, 200] | [50, 100, 200, 500] | [50, 100, 200] | - | - | - |
| Learning Rate | - | [0.01, 0.1, 0.5] | [0.01, 0.1] | [0.01, 0.1, 0.5] | [0.01, 0.1, 0.2] | - | - |
| Max Depth | [1, 5, 7, 15] | [1, 5, 7,10,15] | [1, 5, 7, 10,15] | - | - | - | - |
| Subsample | - | [0.01, 1.0] | [0.01, 1.0] | - | - | - | - |
| Reg Alpha | - | [0, 0.1, 0.5, 1, 2, 3, 5, 10] | [0, 0.1, 0.5, 1, 2, 3, 5, 10] | - | - | - | - |
| Reg Lambda (L2) | - | [0, 0.1, 0.5, 1, 2, 3, 5, 10] | [0, 0.1, 0.5, 1, 2, 3, 5, 10] | - | [0.001, 0.01, 0.1] | - | - |
| Min Samples Leaf | [1, 2, 4] | - | - | - | - | - | - |
| Max features | [0.5, 0.8, 1.0] | - | - | - | - | - | - |
| Min Samples Split | [2, 5, 10] | - | - | - | - | - | - |
| Colsample | - | [0.01, 0.1, 1.0] | [0.01, 0.1, 1.0] | - | - | - | - |
| Dropout Rate | - | - | - | - | [0.2, 0.3, 0.4] | - | - |
| Penalty | - | - | - | - | - | - | ['l1', 'l2'] |
| Criterion | gini | - | - | - | - | - | - |
| C | - | - | - | - | - | [0.1, 1, 10] | [0.1, 1,10] |
| Kernel | - | - | - | - | - | ['linear', 'rbf', 'poly', 'sigmoid'] | - |
| Algorithm | - | - | - | 'SAMME' | - | - | - |
| Gamma | - | 0 | - | - | - | ['scale', 'auto', 0.1, 0.01] | - |
| Solver | - | - | - | - | - | - | ['liblinear', 'lbfgs', 'saga'] |

**Table S8: Comparison of Model Performance between Males and Females.** The performance metrics of the Light Gradient Boosting Machine classifier from the 10-Fold cross-validation are reported as mean ± standard deviation for only males versus only females. Metrics include ROC AUC (area under the receiver operating characteristic curve), PR AUC (area under the precision-recall curve), sensitivity, precision, specificity, accuracy and F1-score.

| Metric | Female | Male |
|---|---|---|
| **Accuracy** | $62.98 \pm 13.63$ | $72.53 \pm 8.84$ |
| **F1 Score** | $58.72 \pm 11.49$ | $70.22 \pm 10.26$ |
| **Precision** | $60.44 \pm 8.95$ | $71.65 \pm 11.63$ |
| **Sensitivity** | $64.56 \pm 13.88$ | $70.84 \pm 9.74$ |
| **Specificity** | $63.19 \pm 24.21$ | $77.26 \pm 13.13$ |
| **PR AUC** | $82.44 \pm 13.34$ | $74.72 \pm 12.31$ |
| **ROC AUC** | $72.84 \pm 8.60$ | $76.71 \pm 10.19$ |

ative agitation events. Finally, the overall scaled mean of each positive or negative condition was calculated per participant.

##### S1.7.1 Poor nighttime sleep and irregular sleep patterns as agitation indicators

The awake state ratio during sleep was calculated as an indicator of the alertness level and number of awakenings during the night. Poor nighttime sleep and increased awake state have been associated with increased sleepiness the following day[13]. Sleep disturbances in dementia have previously been attributed to comorbid sleeping disorders, including disordered breathing, causing apnoea[14]. To explore the correlation of sleep apnoea and poor nighttime sleep in PLwD, the average and minimum nighttime RR were compared between agitation and non-agitation weeks. This analysis aimed to determine whether there was a higher index of sleep apnoea accompanying agitation weeks, which could disrupt sleep and trigger agitation.

The average scaled awake ratio (Figure S10.a) was significantly higher during agitation weeks and the average minimum RR was significantly lower (Figure S10.d) with p values $p-value = 9 \times 10^{-3}$ and $p-value = 2 \times 10^{-2}$ respectively. throughout agitation weeks compared to non-agitation weeks (see Table S14 for statistical analysis results and Table S13 for descriptive statistics).

Since this study investigated a whole week leading up to an agitation event, an analysis of SD allowed us to examine the variability of the measures throughout the week. By analysing weekly SD, significant changes that may have occurred on one day due to agitation could be effectively captured.

We computed the weekly SD for awake ratio and daytime sleep and compared them between agitation and non-agitation weeks. High SD values indicated higher variability and fluctuation throughout the week. To calculate SD, the daily mean values and the weekly mean were used. The SD of each measure was z-score normalised in the same manner as the means.

To further investigate the assumption of fluctuation, Shannon's entropy was calculated[15] to quantify consistency in sleep patterns. It was calculated based on the probability of the mean sleep measure falling into four interval categories. These categories were determined proportionally to the overall mean of the same sleep metric for the given week. The four categories were defined as

**Table S9: Demographics of Consistently Correct and Incorrect Predictions.** Participants with consistently correct and incorrect model predictions across all events are shown with their demographics.

| **Always True (n=19)** | | |
|---|---|---|
| **Diagnosis** | **Female n=8** | **Male n=11** |
| Alzheimer's Disease | 7 | 6 |
| Vascular Dementia | - | 1 |
| Other and mixed | 1 | 1 |
| Unspecified | - | 3 |

| **Always False (n=5)** | | |
|---|---|---|
| **Diagnosis** | **Female n=2** | **Male n=3** |
| Alzheimer's Disease | 1 | 2 |
| Vascular dementia | 1 | - |
| Unspecified | - | 1 |

**Table S10: Estimation of Agitation Frequency.** The agitation frequency reported on the Neuropsychiatric Inventory (NPI) assessments at the baseline time points (Baseline) and on all other time points (Rest) is shown. The number indicates the number of participants within the reported frequency group.

| **Agitation Frequency** | **Baseline** | **Rest** |
|---|---|---|
| Not reported (0) | 84 | 230 |
| Less than once a week (1) | 16 | 71 |
| Once a week (2) | 7 | 46 |
| More than once a week (3) | 7 | 34 |
| Every day (4) | 4 | 9 |
| **Total questionnaires** | **118** | **390** |

follows:

$$\begin{aligned} &\text{Category 1:} && 0.5 \times \text{mean\_week} > \text{mean\_day} \\ &\text{Category 2:} && \text{mean\_week} \geq \text{mean\_day} > 0.5 \times \text{mean\_week} \\ &\text{Category 3:} && 2 \times \text{mean\_week} > \text{mean\_day} \geq \text{mean\_week} \\ &\text{Category 4:} && \text{mean\_day} \geq 2 \times \text{mean\_week} \end{aligned} \tag{2}$$

The daily means were categorised into four intervals, and probabilities were computed for each category based on how often the mean value was assigned to that category. A lower probability reflected a higher entropy and thus higher fluctuations in sleep metrics throughout the week, as the metric values were distributed across multiple categories, rather than consistently belonging to one. The mean weekly entropy was calculated by averaging the daily means for each event. Then, the overall average of the positive or negative condition was calculated per participant.

The average variability in awake ratio and nap duration, as indicated by the mean scaled SD, were found to be significantly higher with p-values $p - value = 4.1 \times 10^{-2}$ and, $p - value = 4.2 \times 10^{-2}$ respectively, during weeks characterised by agitation compared to weeks without agitation (Figure S11). For example, the awake ratio of a participant exhibited greater fluctuations during an agitation week compared to a non-agitation week, with a substantial increase observed on the second day preceding the agitation recording (Figure S11.f). Similarly, the variability in nap duration was higher during agitation weeks compared to non-agitation weeks (Figure S11.c).

Shannon's entropy related to nap duration was significantly higher during agitation weeks compared to non-agitation weeks (Figure S11.c) (see Table S13 for descriptive statistics and Table S14 for the statistical analysis results).

**Table S11: Performance of Light Gradient Boosting Machine (LightGBM) Classifier on Feature Subsets.** Comparison of performance of LightGBM classifiers trained on seven feature subsets each excluding specific categories (Activity, Sleep, Light, and Temperature). Classifier performance comparison is conducted using stratified, ID-grouping 10-Fold cross-validation, using the 20 first selected features from each subset. The performance metrics are reported as mean ± standard deviation. Metrics include ROC AUC (area under the receiver operating characteristic curve), PR AUC (area under the precision-recall curve), sensitivity, precision, specificity, accuracy and F1-score. The row representing the feature subset that yielded the best results is highlighted in bold.

| Features | Accuracy | F1 Score | Precision | Sensitivity | Specificity | PR AUC | ROC AUC |
|---|---|---|---|---|---|---|---|
| **All** | **71.36 ± 7.21** | **71.12 ± 7.28** | **71.81 ± 7.94** | **71.32 ± 7.38** | **75.28 ± 10.43** | **78.60 ± 7.57** | **77.63 ± 6.59** |
| **Act** | 48.44 ± 9.62 | 47.10 ± 10.11 | 49.39 ± 10.91 | 49.43 ± 10.62 | 50.20 ± 19.82 | 51.79 ± 13.08 | 49.74 ± 13.97 |
| **No act** | 67.10 ± 6.42 | 64.85 ± 5.77 | 65.34 ± 5.59 | 66.79 ± 6.89 | 71.80 ± 9.65 | 76.81 ± 10.92 | 76.07 ± 7.25 |
| **No light** | 59.35 ± 6.84 | 58.40 ± 7.38 | 59.26 ± 7.26 | 59.48 ± 7.07 | 61.71 ± 8.80 | 63.11 ± 14.72 | 62.33 ± 12.03 |
| **No sleep** | 61.37 ± 12.89 | 59.05 ± 14.86 | 64.68 ± 7.14 | 65.25 ± 10.50 | 60.18 ± 26.85 | 77.44 ± 15.11 | 74.67 ± 8.39 |
| **No temp** | 54.45 ± 15.82 | 50.93 ± 18.08 | 61.06 ± 9.77 | 58.91 ± 10.02 | 48.03 ± 29.08 | 68.72 ± 14.57 | 66.27 ± 10.49 |
| **Sleep** | 59.85 ± 7.03 | 56.04 ± 6.33 | 56.98 ± 5.58 | 59.84 ± 10.63 | 63.40 ± 19.17 | 59.97 ± 18.60 | 61.70 ± 9.89 |

**Table S12: Measures used in statistical analysis.** Weekly measures used for statistical comparison between agitation and non-agitation weeks

| Category | Measure |
|---|---|
| **Sleep** | Awake Ratio<br>Nap Duration<br>Average Respiratory Rate (RR average)<br>Minimum Respiratory Rate (RR minimum) |
| **Sleep variability / fluctuation** | Awake ratio (SD)<br>Nap duration (SD)<br>Entropy of awake ratio<br>Entropy of nap duration |
| **Indoor Light Exposure and Indoor Temperature** | **Morning**<br>Lounge<br>Kitchen<br>Bedroom<br>Bathroom<br>**Afternoon**<br>Lounge<br>Kitchen<br>Bedroom<br>Bathroom<br>**Evening**<br>Lounge<br>Kitchen<br>Bedroom<br>Bathroom<br>**Night**<br>Lounge<br>Kitchen<br>Bedroom<br>Bathroom |
| **Ambient Quality-Related measures** | Overall indoor illuminance<br>Outdoor Illuminance<br>Duration of outdoor light exposure<br>Ratio (Indoor:Outdoor Illuminance)<br>Overall indoor temperature<br>Outdoor Temperature<br>Temperature Ratio (Indoor:Outdoor Temperature) |

**Table S13: Descriptive statistics for Sleep Measures.** (Mean(±SD)) and Scaled Mean(±SD) for the comparison of the different sleep measures between agitation and non-agitation weeks.

| Variable | State | Mean ± SD | Scaled Mean ± SD |
|---|---|---|---|
| Awake ratio | Agitated | 0.24±0.16 | 0.27±0.67 |
| | Non-agitated | 0.22±0.17 | -0.18±0.40 |
| Nap duration | Agitated | 0.32±0.54 | 0.18±0.46 |
| | Non-agitated | 0.17±0.26 | -0.14±0.32 |
| Average respiratory rate | Agitated | 14.41±1.76 | -0.06±0.64 |
| | Non-agitated | 14.52±1.68 | 0.09±0.43 |
| Minimum respiratory rate | Agitated | 9.00±0.66 | -0.33±0.68 |
| | Non-agitated | 9.13±0.55 | 0.09±0.33 |
| Awake ratio variability (SD) | Agitated | 0.24±0.16 | 0.21±0.59 |
| | Non-agitated | 0.22±0.17 | -0.17±0.43 |
| Awake ratio entropy | Agitated | 0.86±0.16 | 0.02±0.48 |
| | Non-agitated | 0.84±0.16 | -0.07±0.39 |
| Nap duration variability (SD) | Agitated | 0.25±0.48 | 0.23±0.48 |
| | Non-agitated | 0.13±0.24 | -0.18±0.35 |
| Entropy nap duration | Agitated | 0.36±0.35 | 0.25±0.59 |
| | Non-agitated | 0.24±0.23 | -0.14±0.36 |

**Table S14: Statistical results for the Comparison of Sleep Measures between agitation and non-agitation weeks** Results of paired T-tests for comparison of sleep measures between agitation and non-agitation weeks. The uncorrected p-values are reported. Significant results ($p-value < 0.05$) are highlighted in bold.

| Variable | DDOF | T-statistic | p-value | CI95% | Cohen-D | Power |
|---|---|---|---|---|---|---|
| **Awake ratio** | **1,28** | **2.77** | **0.009** | **[0.02, 0.16]** | **0.82** | **0.99** |
| Nap duration | 1,21 | 2.10 | 0.05 | [0.0, 0.64] | 0.80 | 0.95 |
| Average respiratory rate | 1,28 | 0.84 | 0.41 | [-0.5,0.21] | 0.27 | 0.28 |
| **Minimum respiratory rate** | **1,28** | **-2.57** | **0.02** | **[-0.76, -0.09]** | **0.79** | **0.98** |
| **Awake ratio variability (SD)** | **1,28** | **2.35** | **0.03** | **[0.05, 0.71]** | **0.74** | **0.97** |
| Awake ratio entropy | 1,28 | 0.58 | 0.57 | [-0.22, 0.39] | 0.19 | 0.17 |
| **Nap duration variability (SD)** | **1,21** | **2.60** | **0.02** | **[0.08, 0.72]** | **0.96** | **0.99** |
| **Entropy nap duration** | **1,21** | **2.17** | **0.04** | [0.02, 0.77] | **0.80** | **0.95** |

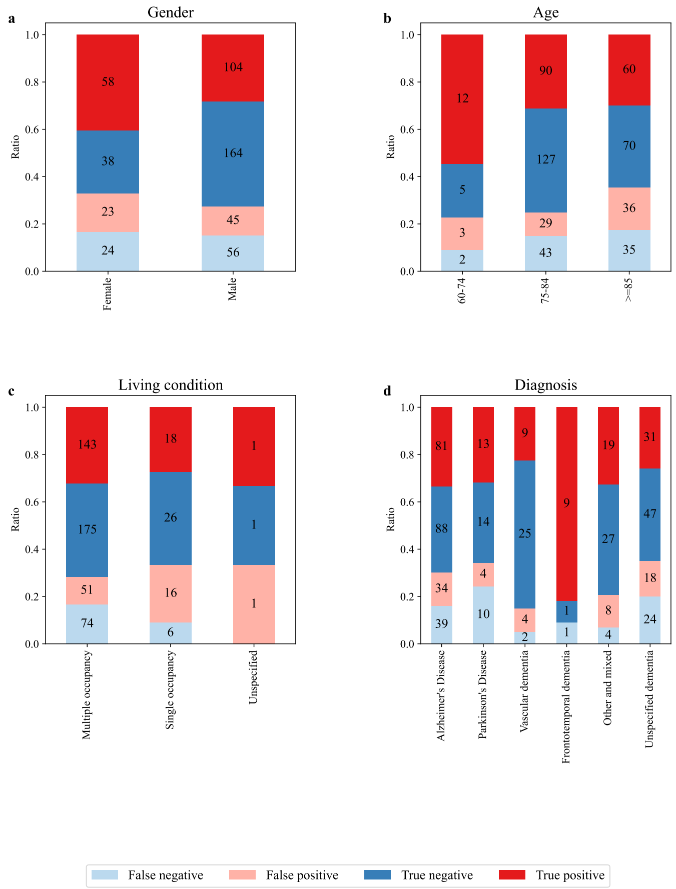


**Figure S2: Bias analysis.** For each demographic group, the ratio of model prediction outcomes [True Positives (TP), True Negatives (TN), False Positives (FP), False Negatives (FN)] are shown. The size of the bar portion indicates the ratio, with annotated numbers within each bar fraction representing the actual count. Blue: TN, Red: TP, Light blue: FN, Pink: FP. (a) Across Gender (b) Age groups (c) Living condition (d) Dementia type

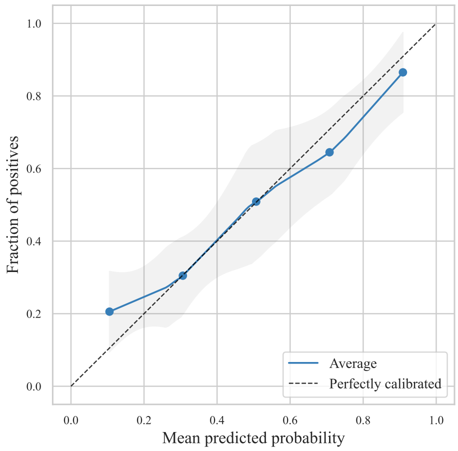


**Figure S3: Calibration plot.** The average agitation-predicted risk is plotted against the proportion of positive agitation episodes. This was calculated on the stratified, ID-grouping 10-Fold evaluation of the Light Gradient Boosting Machine classifier. The grey shaded area shows the standard deviation (SD) across the 10 Folds.

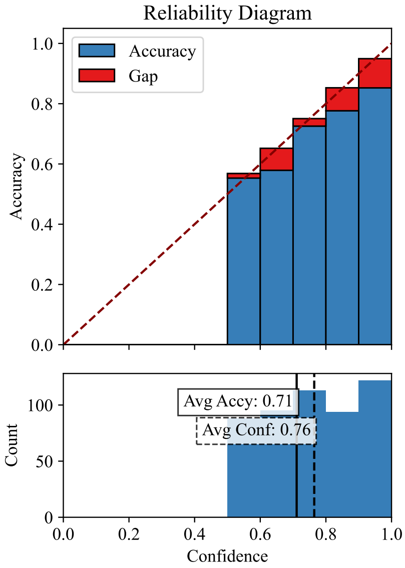


**Figure S4: Reliability plot.** [Top] The model confidence (on positive and negative agitation cases) is shown against accuracy on the test sets from the stratified 10-Fold, ID-grouping evaluations. The gap shows the difference between the average accuracy and confidence of a bin, which would ideally be 0. [Bottom] The histogram of confidences is shown as reported by the model on the test sets.

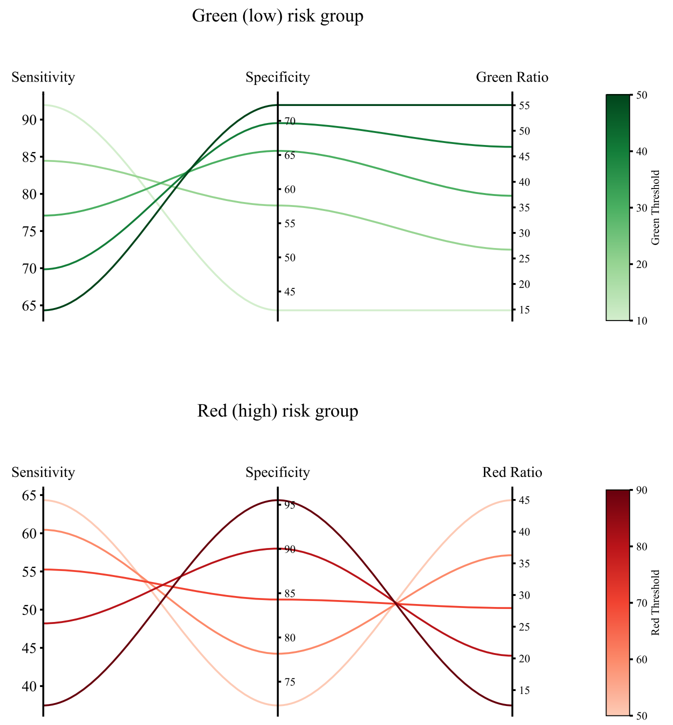


**Figure S5: Determination of Risk Stratification Thresholds.** The variations in sensitivity and specificity on the validation sets are shown when choosing different thresholds. The line colours represent different threshold values, which increase with a resolution of 10% while evaluating the performance on the validation sets within the nested-10-fold validation using only the training set of the external validation. The sensitivity, specificity, and ratios are averaged for each threshold across the 10 folds and are calculated while only considering the Red and Green alerts. For the purpose of the visualisation for the Green threshold figure the Red threshold was set at $> 50\%$, and for the Red group figure, the Green threshold was set at $\leq 50\%$. However, all combinations of thresholds were explored during the analysis.

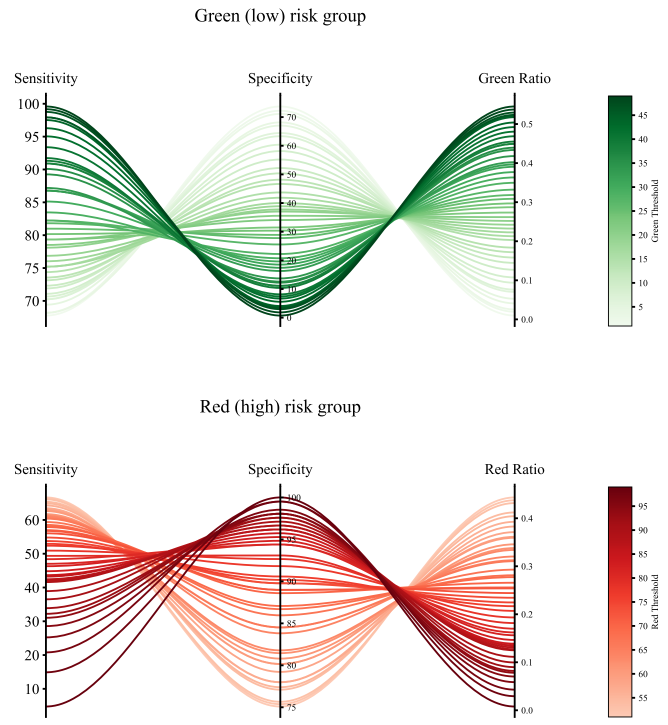


**Figure S6: Effect of Risk Stratification** The variations in sensitivity and specificity on the test sets are shown when different thresholds are chosen. The line colours represent different threshold values. The thresholds increase with a resolution of 1% for the purposes of the visualisation. Sensitivity and specificity are calculated on the test sets corresponding to Red and Green alerts. To calculate the metrics on the Green group, the Red threshold was set at $> 50\%$, and when calculating the metrics on the Red group, the Green threshold was set at $\leq 50\%$.

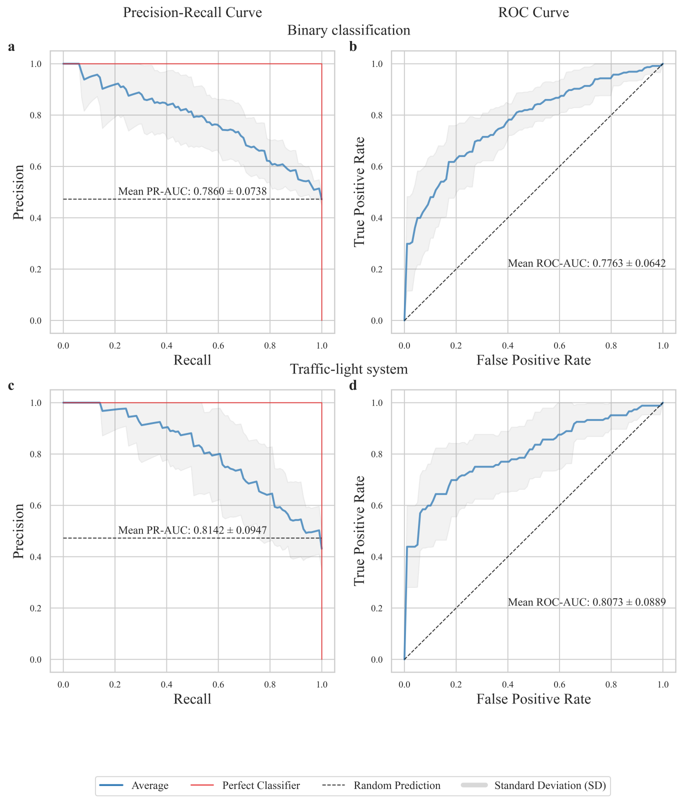


**Figure S7: Precision-Recall (PR) and Receiver-Operating Characteristic (ROC) Curves for Gradient Boosting (LightGBM) binary classification and classification after traffic-light based stratification.** The blue lines represent the average Area Under the Curve (AUC) scores over the 10 Folds. The grey area represents the standard deviation (SD) across the 10 Folds. (a) PR Curve for binary classification. (b) ROC Curve for binary classification. (c) PR Curve for traffic-light-based classification. (d) ROC Curve for traffic-light-based classification.

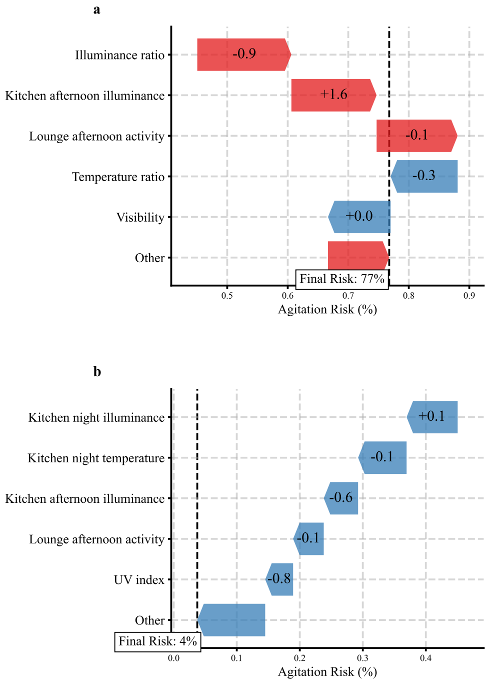


**Figure S8: Personalised investigation of modifiable features.** Examples from a positive (a) and a negative (b) event from PLwD are shown using the SHAP framework. The colour of the arrow corresponds to the contribution: red contributes to agitation presence and blue contributes to agitation absence. Positive SHAP values contributed to positive predictions (agitated), while negative SHAP values contributed to negative predictions (non-agitated). The size of the arrow represents the absolute SHAP value, indicating the magnitude of each feature's contribution. The number within the arrow corresponds to the normalised feature value.

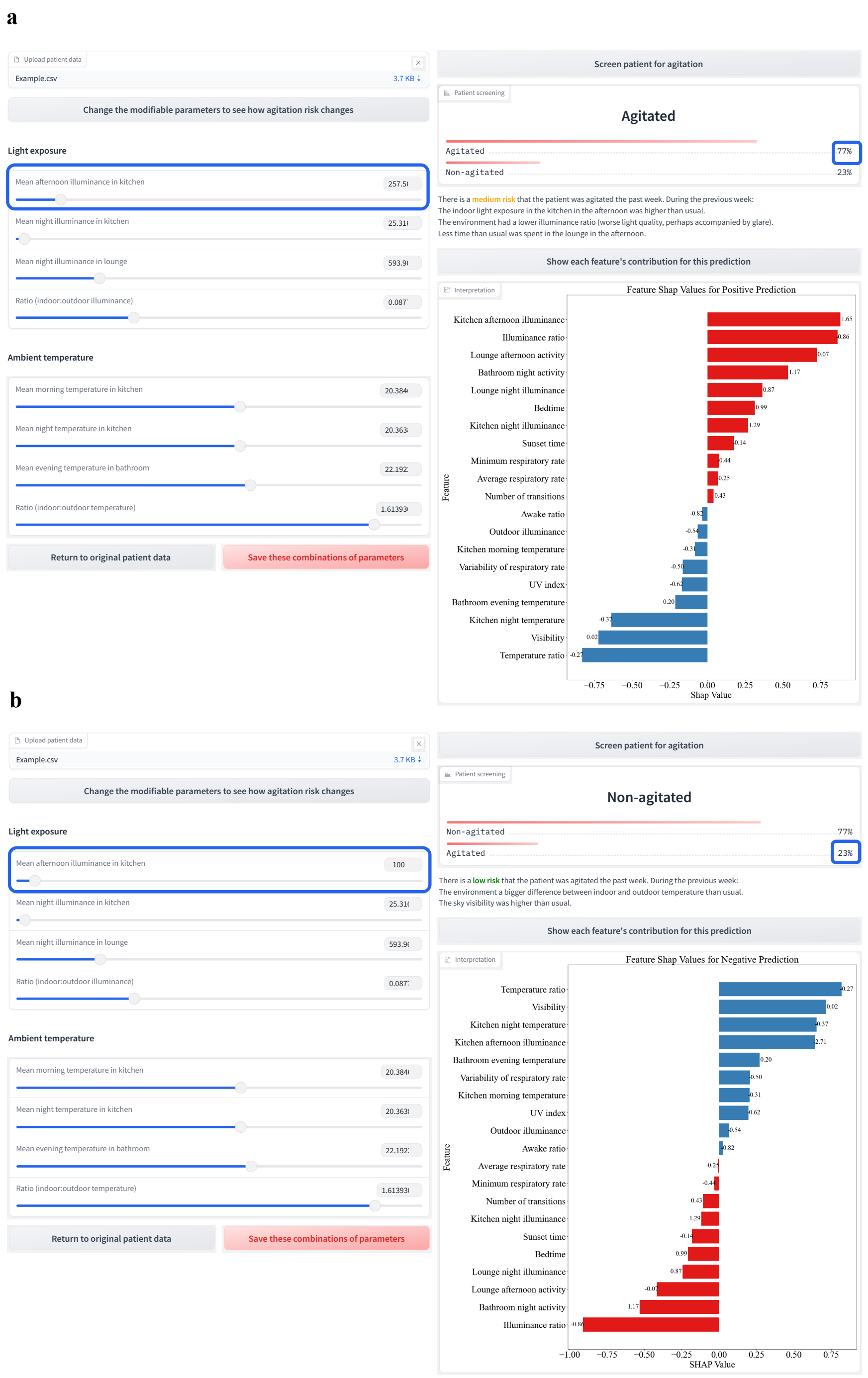


**Figure S9: *In-silico* Experiment: Adjusting Light Exposure via an Interactive Interface** The interactive interface is shown, which accepts the input data as a CSV file. The tool provides sliding bars for the modifiable features and presents the predicted risk and associated probabilities, and a feature importance plot using SHAP values. In the feature importance plot, red bars correspond to features that contributed towards positive agitation prediction and blue bars correspond to features that predict the absence of agitation. Each bar is annotated with the corresponding normalised feature value. The user can save the combinations of modifications they have made to the modifiable parameters. a) Anonymised data from a participant. b) The results after modifying one of the parameters, morning indoor illuminance in the kitchen. An online version with a synthetic patient data generator is hosted on huggingface. (see `https://huggingface.co/spaces/marirena/AgitationScreening`). Access to the tool is currently restricted. Upon publication of our work, access will become open.

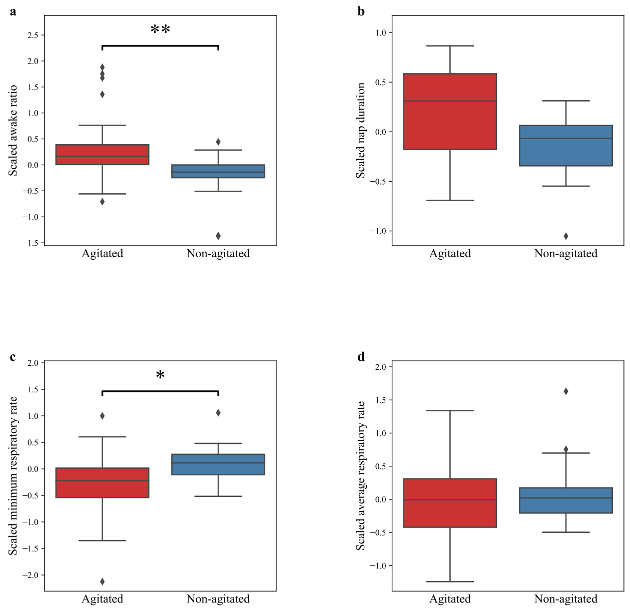


**Figure S10: Sleep measures comparison between agitation and non-agitation weeks.** The boxplots depict the median and the quartiles, with the whiskers showing the 1.5 interquartile range. Significant differences are shown with asterisks. (a) Mean awake ratio (b) Mean nap duration (c) Mean minimum respiratory rate (RR) (d) Mean average respiratory rate (RR).

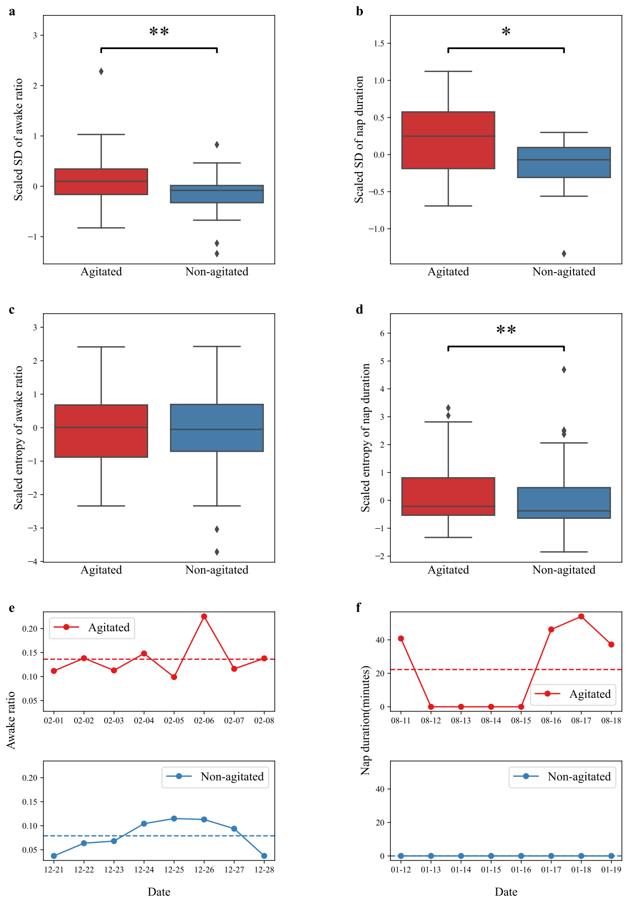


**Figure S11: Sleep measures variability comparison between agitation and non-agitation weeks.** (a),(d) Mean variability (SD) of (a) awake ratio, (d) nap duration, averaged per event, scaled across each participant's events, averaged for positive/negative condition per participant. (b),(e) Shannon's entropy of probabilities of (b) mean awake state and (e) nap duration. (c),(f) Examples from participant B. (c) Fluctuation of awake ratio (f) Fluctuation of nap duration, over the 8 days preceding the agitation event (above) and non-agitation event (below).

###### S1.7.2 Increased indoor lighting and poor light quality as indicators of increased agitation risk

An investigation of the effect of indoor illuminance on agitation found that (average, scaled per participant, location, time-period) illuminance was significantly higher during agitation weeks (Mean(±SD) = 0.10(±0.25)) than non-agitation weeks (Mean(±SD) = -0.05(±0.19), t(1,28) = 2.135, $p-value = 4\times10^{-2}$, Cohen-d = 0.66). A more detailed comparison investigated light exposure differences across different rooms and time-periods between agitation and non-agitation weeks. Figure S12 displays the unprocessed illuminance values, indicating increased illuminance during the daytime in agitation weeks and similar illuminance during the night, except for the lounge area. Significant differences in illuminance existed in the morning in the lounge ($p-value = 3\times10^{-2}$) and in the afternoon in the kitchen ($p-value = 4\times10^{-2}$) (see Table S15 for descriptive statistics and Table S16 for statistical analysis results). To further investigate the effect of light exposure on agitation, outdoor illuminance was compared between agitation and non-agitation weeks. The outdoor illuminance values (Table S21.c) were significantly higher ($p-value = 10^{-2}$) during agitation weeks compared to non-agitation weeks (see Table S15 for descriptive statistics and Table S16 for the statistical analysis results).

Since indoor light sensors cannot distinguish indoor illuminance, generated by light bulbs, and outdoor illuminance coming through the windows, it was necessary to assess whether the observed higher indoor illuminance resulted exclusively from high outdoor illuminance. This could influence potential strategies designed to prevent and manage agitation events.

To account for the influence of sky clarity on indoor illuminance values, the days were separated based on sky conditions. Clear-sky days exhibit higher illuminance levels, which can impact indoor illuminance, while overcast days are characterised by lower illuminance. Each day within labelled weeks was treated as a separate agitation or non-agitation event due to changing weather conditions between days. Out of 29 participants, only 18 had confirmed labels of both positive and negative agitation events for all three types of sky conditions. For these participants, a two-way repeated-measures Analysis Of Variance (ANOVA) was conducted.

The values of indoor illuminance were significantly higher ($p-value = 2\times10^{-2}$) in instances of agitation for all different types of sky (Figure S13.a). There was no interaction between sky clarity and agitation ($p-value > 5\times10^{-2}$) (see Table S15 for descriptive statistics and Table S16 for the statistical analysis results).

Since we showed that higher indoor illuminance was not attributed to higher outdoor illuminance during agitation weeks (Figure S13.b), it was important to further investigate the relation between outdoor and indoor light, to understand whether their interaction could have affected the light quality. For this reason, the ratio of indoor to outdoor illuminance was calculated. This ratio can capture how well-lit a room is[16]. The ratio was significantly higher during non-agitation weeks compared to agitation weeks ($p-value < 10^{-4}$) (Figure S13.c) (see Table S17 for descriptive statistics and Table S18, Table S21 for the statistical analysis results).

**Table S15: Descriptive Statistics for the Comparison of Indoor Illuminance Measures for Agitation and non-agitation weeks.** (Mean(±SD)) and Scaled Mean(±SD) for the comparison of scaled indoor illuminance in different rooms and time-period combinations between agitation and non-agitation weeks.

| Time-period | Location | State | Mean ± SD | Scaled Mean ± SD |
|---|---|---|---|---|
| Morning | | | | |
| | Lounge | Agitated | $345.90 \pm 319.81$ | $0.17 \pm 0.42$ |
| | | Non-agitated | $303.39 \pm 240.90$ | $-0.10 \pm 0.30$ |
| | Kitchen | Agitated | $343.81 \pm 240.90$ | $0.24 \pm 0.60$ |
| | | Non-agitated | $281.15 \pm 145.12$ | $-0.09 \pm 0.28$ |
| | Bedroom | Agitated | $275.64 \pm 184.78$ | $0.10 \pm 0.37$ |
| | | Non-agitated | $270.63 \pm 242.85$ | $-0.06 \pm 0.27$ |
| | Bathroom | Agitated | $254.84 \pm 137.38$ | $0.16 \pm 0.56$ |
| | | Non-agitated | $229.98 \pm 117.99$ | $-0.08 \pm 0.32$ |
| Afternoon | | | | |
| | Lounge | Agitated | $347.69 \pm 339.68$ | $0.18 \pm 0.46$ |
| | | Non-agitated | $299.72 \pm 225.61$ | $-0.07 \pm 0.40$ |
| | Kitchen | Agitated | $455.18 \pm 418.60$ | $0.22 \pm 0.58$ |
| | | Non-agitated | $327.50 \pm 244.87$ | $-0.11 \pm 0.30$ |
| | Bedroom | Agitated | $327.07 \pm 266.84$ | $0.15 \pm 0.44$ |
| | | Non-agitated | $309.04 \pm 287.00$ | $-0.04 \pm 0.26$ |
| | Bathroom | Agitated | $312.04 \pm 233.00$ | $0.12 \pm 0.63$ |
| | | Non-agitated | $267.17 \pm 164.95$ | $-0.04 \pm 0.33$ |
| Evening | | | | |
| | Lounge | Agitated | $98.18 \pm 124.90$ | $-0.06 \pm 0.39$ |
| | | Non-agitated | $104.09 \pm 129.31$ | $0.01 \pm 0.29$ |
| | Kitchen | Agitated | $95.86 \pm 57.33$ | $0.10 \pm 0.66$ |
| | | Non-agitated | $83.02 \pm 63.10$ | $-0.07 \pm 0.33$ |
| | Bedroom | Agitated | $66.89 \pm 77.04$ | $-0.02 \pm 0.52$ |
| | | Non-agitated | $67.64 \pm 73.08$ | $0.08 \pm 0.47$ |
| | Bathroom | Agitated | $159.09 \pm 130.82$ | $0.05 \pm 0.67$ |
| | | Non-agitated | $147.56 \pm 118.68$ | $0.03 \pm 0.55$ |
| Night | | | | |
| | Lounge | Agitated | $94.29 \pm 107.79$ | $0.08 \pm 0.51$ |
| | | Non-agitated | $76.48 \pm 110.80$ | $-0.08 \pm 0.34$ |
| | Kitchen | Agitated | $56.47 \pm 57.85$ | $0.15 \pm 0.58$ |
| | | Non-agitated | $47.95 \pm 64.25$ | $-0.22 \pm 0.35$ |
| | Bedroom | Agitated | $38.28 \pm 39.71$ | $-0.03 \pm 0.38$ |
| | | Non-agitated | $44.84 \pm 78.13$ | $-0.03 \pm 0.37$ |
| | Bathroom | Agitated | $133.78 \pm 140.33$ | $0.02 \pm 0.53$ |
| | | Non-agitated | $134.04 \pm 146.78$ | $0.01 \pm 0.40$ |

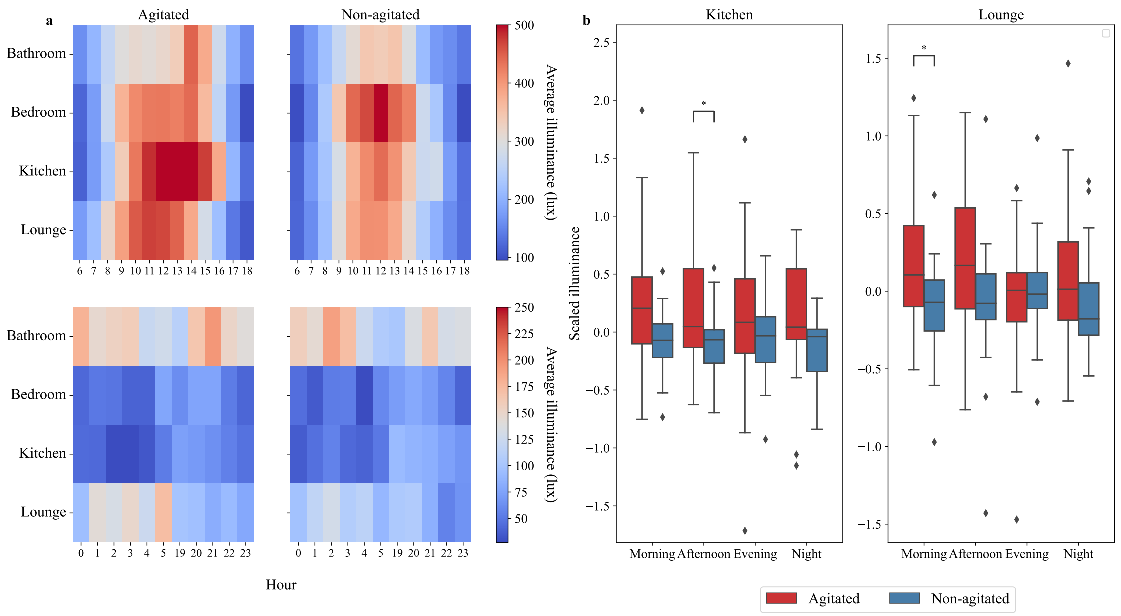


**Figure S12: Light Exposure Measures Comparison between agitation and non-agitation weeks** (a) Unprocessed values of mean illuminance averaged across all participants, per room and hour 6 a.m.-6 p.m. (above) and 6 p.m.-6 a.m. (below) for agitation and non-agitation weeks. (b) Average scaled (per participant, location, time-period) illuminance in different rooms and time-periods for positive and non-negative conditions.

**Table S16: Statistical Results for the Comparison of Indoor Illuminance Measures between agitation and non-agitation weeks.** Results of paired T-tests for comparison of scaled indoor illuminance between agitation and non-agitation weeks. The first row represents the average indoor illuminance across rooms and time-periods. All other rows represent one combination of a room and time-period each. The uncorrected p-values are reported. Note: bold if $p - value < 0.05$.

| Variable | Room | DDOF | T-statistic | p-value | CI95% | Cohen-D | Power |
|---|---|---|---|---|---|---|---|
| **Average indoor illuminance** | - | **1,28** | **2.14** | **0.04** | **[0.01, 0.29]** | **0.66** | **0.93** |
| Morning | **Lounge** | **1,28** | **2.28** | **0.03** | **[0.03, 0.51]** | **0.74** | **0.97** |
| | Kitchen | 1,25 | 2.09 | 0.05 | [0.00, 0.66] | 0.70 | 0.93 |
| | Bedroom | 1,27 | 1.48 | 0.15 | [-0.06, 0.38] | 0.49 | 0.71 |
| | Bathroom | 1,28 | 1.58 | 0.13 | [-0.07, 0.54] | 0.51 | 0.76 |
| Afternoon | Lounge | 1,28 | 1.80 | 0.08 | [-0.03, 0.53] | 0.57 | 0.85 |
| | **Kitchen** | **1,26** | **2.12** | **0.04** | **[0.01, 0.64]** | **0.70** | **0.94** |
| | Bedroom | 1,27 | 1.56 | 0.13 | [-0.06, 0.45] | 0.53 | 0.77 |
| | Bathroom | 1,28 | 0.98 | 0.33 | [-0.17, 0.47] | 0.31 | 0.36 |
| Evening | Lounge | 1,28 | -0.58 | 0.56 | [-0.29, 0.16] | 0.19 | 0.16 |
| | Kitchen | 1,26 | 1.01 | 0.32 | [-0.19, 0.52] | 0.33 | 0.38 |
| | Bedroom | 1,27 | -0.59 | 0.56 | [-0.41,0.23] | 0.19 | 0.16 |
| | Bathroom | 1,28 | 0.11 | 0.91 | [-0.38, 0.42] | 0.04 | 0.05 |
| Night | Lounge | 1,21 | 0.95 | 0.35 | [-0.19, 0.50] | 0.36 | 0.36 |
| | Kitchen | 1,18 | 1.82 | 0.09 | [-0.06, 0.79] | 0.77 | 0.88 |
| | Bedroom | 1,27 | -0.04 | 0.97 | [-0.27, 0.26] | 0.01 | 0.05 |
| | Bathroom | 1,26 | 0.09 | 0.93 | [-0.31, 0.33] | 0.03 | 0.05 |

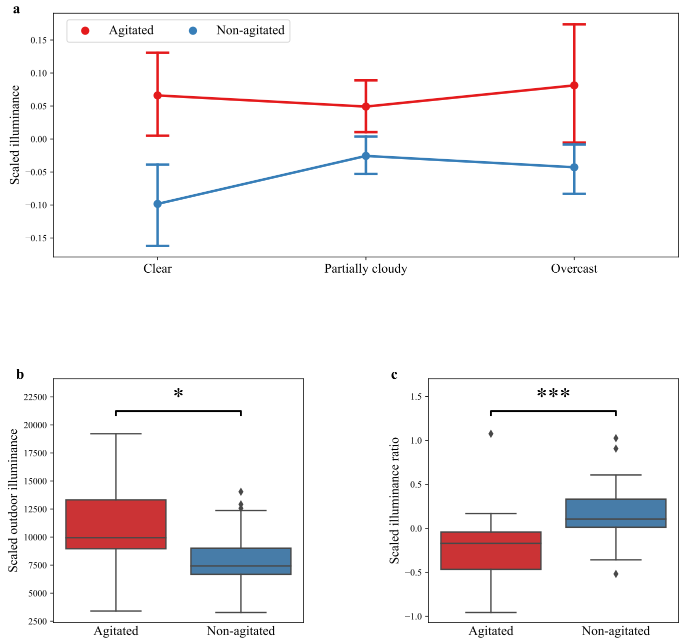


**Figure S13: Comparison of the Combination of Outdoor and Indoor Light Exposure Measures between agitation and non-agitation weeks.** (a) Each date was considered an agitation or non-agitation event. Indoor illuminance values were scaled per participant, location, time-period, and type of sky. The point plot shows the difference in illuminance between agitation and non-agitation days for three different sky clarity types (clear, partially cloudy, and overcast). Error bars show a 95% confidence interval for each sky type, for each condition. (b) Average scaled (across all participants) outdoor illuminance for positive and negative conditions. (c) Average scaled (per participant) ratio of indoor (morning & afternoon) illuminance: outdoor (daylight) illuminance between positive and negative conditions.

**Table S17: Descriptive Statistics for the Comparison of Outdoor Illuminance Related Measures between agitation and non agitation weeks.** Mean±SD of different variables related to indoor and outdoor light exposure, for comparison between agitation and non-agitation weeks.

| Variable | State | Mean ± SD | Scaled Mean ± SD |
|---|---|---|---|
| Overall indoor illuminance (lux) | Agitated | 216.14 ± 79.03 | 0.10 ± 0.25 |
| | Non-agitated | 195.12 ± 78.73 | -0.05 ± 0.19 |
| Outdoor illuminance (lux) | Agitated | 9833.05 ± 3878.90 | 0.25 ± 0.63 |
| | Non-agitated | 7226.55 ± 2644.50 | -0.20 ± 0.33 |
| Exposure duration (minutes) | Agitated | 199.51 ± 74.87 | -0.16 ± 0.44 |
| | Non-agitated | 218.19 ± 78.33 | 0.02 ± 0.40 |
| Clear days indoor illuminance | Agitated | 450.87 ± 240.90 | 0.07 ± 0.21 |
| | Non-agitated | 368.35 ± 181.89 | -0.11 ± 0.21 |
| Cloudy days indoor illuminance | Agitated | 287.87 ± 120.80 | 0.05 ± 0.13 |
| | Non-agitated | 267.29 ± 100.49 | -0.03 ± 0.09 |
| Overcast days indoor illuminance | Agitated | 146.02 ± 72.32 | 0.09 ± 0.26 |
| | Non-agitated | 132.57 ± 99.24 | -0.05 ± 0.13 |
| Illuminance ratio | Agitated | 0.04 ± 0.002 | -0.26 ± 0.27 |
| | Non-agitated | 0.05 ± 0.03 | 0.18 ± 0.26 |
| Overall indoor temperature | Agitated | 21.08 ± 1.59 | -0.01± 0.45 |
| | Non-agitated | 21.21 ±1.30 | 0.08± 0.28 |
| Outdoor temperature | Agitated | 11.71 ± 3.50 | 0.00 ± 0.68 |
| | Non-agitated | 11.63 ± 2.33 | -0.04 ± 0.29 |
| Temperature ratio | Agitated | 2.56 ± 0.99 | -0.04± 0.35 |
| | Non-agitated | 2.83 ± 0.92 | 0.01 ± 0.24 |

**Table S18: Statistical Results for the Comparison of Indoor Illuminance Measures between agitation and non-agitation days.** Results of Two-Way Repeated-measures ANOVA for comparison of scaled indoor illuminance for different types of sky clarity between agitation and non-agitation days. The uncorrected p-values are reported. Note: bold if $p - value < 0.05$. **SS**: Sum of Squares **MS**: Mean of Squares, **ng**$^2$: Generalised eta-squared effect size

| Variable | SS | DDOF | MS | p-value | F-statistic | ng$^2$ |
|---|---|---|---|---|---|---|
| Agitation state | **0.50** | **1,18** | **0.49** | **0.02** | **6.06** | **0.13** |
| Sky clarity | 0.05 | 1,18 | 0.02 | 0.11 | 2.37 | 0.01 |
| Agitation state × Sky clarity | 0.08 | 1,18 | 0.02 | 0.50 | 0.71 | 0.01 |

**Table S19: Descriptive Statistics for the Comparison of Indoor Temperature Measures between agitation and non-agitation weeks.** (Mean(±SD)) and Scaled Mean(±SD) for the comparison of scaled indoor temperature in different rooms and time-period combinations between agitation and non-agitation weeks.

| Time-period | Location | State | Mean ± SD | Scaled Mean ± SD |
|---|---|---|---|---|
| Morning | | | | |
| | Lounge | Agitated | $20.86 \pm 1.38$ | $-0.00 \pm 0.53$ |
| | | Non-agitated | $21.18 \pm 1.44$ | $0.11 \pm 0.32$ |
| | Kitchen | Agitated | $20.87 \pm 1.95$ | $0.00 \pm 0.60$ |
| | | Non-agitated | $20.82 \pm 1.29$ | $0.07 \pm 0.27$ |
| | Bedroom | Agitated | $20.50 \pm 1.68$ | $-0.00 \pm 0.57$ |
| | | Non-agitated | $20.92 \pm 1.77$ | $0.09 \pm 0.34$ |
| | Bathroom | Agitated | $20.49 \pm 2.10$ | $-0.13 \pm 0.43$ |
| | | Non-agitated | $20.79 \pm 1.32$ | $0.10 \pm 0.27$ |
| Afternoon | | | | |
| | Lounge | Agitated | $21.39 \pm 1.38$ | $0.02 \pm 0.55$ |
| | | Non-agitated | $21.88 \pm 1.33$ | $0.06 \pm 0.32$ |
| | Kitchen | Agitated | $22.11 \pm 1.81$ | $0.05 \pm 0.61$ |
| | | Non-agitated | $21.82 \pm 1.67$ | $0.08 \pm 0.33$ |
| | Bedroom | Agitated | $21.33 \pm 1.31$ | $0.07 \pm 0.57$ |
| | | Non-agitated | $21.27 \pm 1.65$ | $0.04 \pm 0.33$ |
| | Bathroom | Agitated | $21.49 \pm 1.58$ | $-0.03 \pm 0.41$ |
| | | Non-agitated | $21.38 \pm 1.5$ | $0.06 \pm 0.31$ |
| Evening | | | | |
| | Lounge | Agitated | $21.53 \pm 1.54$ | $0.02 \pm 0.54$ |
| | | Non-agitated | $21.85 \pm 1.69$ | $0.11 \pm 0.36$ |
| | Kitchen | Agitated | $21.88 \pm 1.83$ | $0.15 \pm 0.59$ |
| | | Non-agitated | $21.48 \pm 1.45$ | $0.01 \pm 0.32$ |
| | Bedroom | Agitated | $21.36 \pm 1.66$ | $0.09 \pm 0.59$ |
| | | Non-agitated | $21.68 \pm 1.67$ | $0.05 \pm 0.36$ |
| | Bathroom | Agitated | $21.41 \pm 2.14$ | $-0.17 \pm 0.38$ |
| | | Non-agitated | $21.59 \pm 1.25$ | $0.13 \pm 0.32$ |
| Night | | | | |
| | Lounge | Agitated | $20.62 \pm 1.32$ | $-0.02 \pm 0.48$ |
| | | Non-agitated | $20.88 \pm 1.58$ | $0.08 \pm 0.35$ |
| | Kitchen | Agitated | $20.86 \pm 1.69$ | $0.10 \pm 0.57$ |
| | | Non-agitated | $20.61 \pm 1.30$ | $-0.03 \pm 0.27$ |
| | Bedroom | Agitated | $20.09 \pm 1.66$ | $0.03 \pm 0.58$ |
| | | Non-agitated | $20.33 \pm 1.74$ | $0.08 \pm 0.35$ |
| | Bathroom | Agitated | $20.08 \pm 2.07$ | $-0.14 \pm 0.42$ |
| | | Non-agitated | $20.37 \pm 1.45$ | $0.14 \pm 0.36$ |

**Table S20: Statistical Results for the Comparison of Indoor Temperature Measures between agitation and non-agitation weeks.** Results of paired T-tests for comparison of scaled indoor temperature between agitation and non-agitation weeks. The first row represents the average indoor temperature across rooms and time-periods. All other rows represent one combination of a room and time-period each. The uncorrected p-values are reported. Note: bold if $p$-value $< 0.05$.

| Time-period | Room | DDOF | T-statistic | p-value | CI95% | Cohen-D | Power |
|---|---|---|---|---|---|---|---|
| Average indoor temperature | - | 1,28 | -0.60 | 0.55 | [-0.27, 0.15] | 0.19 | 0.17 |
| Morning | Lounge | 1,27 | -0.81 | 0.43 | [-0.41, 0.18] | 0.26 | 0.26 |
| | Kitchen | 1,24 | -0.42 | 0.67 | [-0.39, 0.26] | 0.15 | 0.11 |
| | Bedroom | 1,27 | -0.61 | 0.55 | [-0.41, 0.22] | 0.20 | 0.17 |
| | Bathroom | 1,28 | -1.97 | 0.06 | [-0.47, 0.01] | 0.64 | 0.91 |
| Afternoon | Lounge | 1,27 | -0.26 | 0.79 | [-0.33, 0.26] | 0.08 | 0.07 |
| | Kitchen | 1,25 | -0.18 | 0.86 | [-0.37, 0.31] | 0.06 | 0.06 |
| | Bedroom | 1,27 | 0.24 | 0.81 | [-0.27, 0.34] | 0.08 | 0.07 |
| | Bathroom | 1,28 | -0.78 | 0.44 | [-0.34, 0.15] | 0.26 | 0.27 |
| Evening | Lounge | 1,27 | -0.55 | 0.59 | [-0.39, 0.22] | 0.18 | 0.15 |
| | Kitchen | 1,25 | 0.88 | 0.39 | [-0.19,0.47] | 0.30 | 0.31 |
| | Bedroom | 1,27 | 0.23 | 0.82 | [-0.29, 0.37] | 0.08 | 0.07 |
| | **Bathroom** | **1,28** | **-2.66** | **0.01** | **[-0.53, -0.07]** | **0.85** | **0.99** |
| Night | Lounge | 1,24 | -0.68 | 0.50 | [-0.40, 0.20] | 0.23 | 0.20 |
| | Kitchen | 1,20 | 0.76 | 0.45 | [-0.23, 0.49] | 0.29 | 0.25 |
| | Bedroom | 1,27 | -0.37 | 0.71 | [-0.38, 0.26] | 0.12 | 0.10 |
| | **Bathroom** | **1,28** | **-2.28** | **0.03** | **[-0.53, -0.03]** | **0.72** | **0.96** |

**Table S21: Statistical Results for the Comparison of Outdoor Ambient Related Measures between agitation and non-agitation weeks.** Results of paired T-tests for comparison of scaled outdoor illuminance, scaled duration of exposure, scaled illuminance ratio, scaled outdoor temperature and scaled temperature ratio between agitation and non-agitation weeks. The uncorrected p-values are reported. Note: bold if $p - value < 0.05$.

| Variable | DDOF | T-statistic | p-value | CI95% | Cohen-D | Power |
|---|---|---|---|---|---|---|
| **Outdoor illuminance** | **1,23** | **2.65** | **0.01** | **[0.10, 0.80]** | **0.89** | **0.99** |
| Exposure duration | 1,23 | -1.25 | 0.23 | [-0.49, 0.12] | 0.44 | 0.54 |
| **Illuminance Ratio** | **1,28** | **-5.51** | **0.000** | **[-0.60, -0.27]** | **1.65** | **1.0** |
| Outdoor temperature | 1,23 | 0.27 | 0.79 | [-0.32, 0.42] | 0.09 | 0.07 |
| Temperature Ratio | 1,28 | -0.54 | 0.60 | [-0.27, 0.16] | 0.19 | 016 |

#### References


1. Ream, E. *MINDER Health Management Study for Dementia: A study to refine and evaluate technologies in the home to monitor and manage the health of people with dementia* Oct. 10, 2018. http://www.isrctn.com/ISRCTN71000991.

2. Mckinney, W. pandas: a Foundational Python Library for Data Analysis and Statistics. *Python High Performance Science Computer.* 2011.

3. Van Der Walt, S., Colbert, S. C. & Varoquaux, G. The NumPy Array: A Structure for Efficient Numerical Computation. *Computing in Science & Engineering.* 2011. **13.** 22–30. doi:10.1109/MCSE.2011.37.

4. Virtanen, P., Travis E. Oliphant & Ralf Gommers et al. SciPy 1.0: fundamental algorithms for scientific computing in Python. *Nature Methods.* 2020. **17.** 261–272. doi:10.1038/s41592-019-0686-2.

5. Pedregosa, F., Varoquaux, G., Gramfort, A. & et al. *Scikit-learn: Machine Learning in Python* June 5, 2018. arXiv: 1201.0490[cs]. http://arxiv.org/abs/1201.0490.

6. TensorFlow Developers. *TensorFlow* version v2.12.1. July 5, 2023. https://zenodo.org/record/4724125.

7. Waskom, M. seaborn: statistical data visualization. *Journal of Open Source Software.* 2021. **6.** 3021. doi:10.21105/joss.03021.

8. Hunter, J. D. Matplotlib: A 2D Graphics Environment. *Computing in Science & Engineering.* 2007. **9.** 90–95. doi:10.1109/MCSE.2007.55.

9. Vallat, R. Pingouin: statistics in Python. *Journal of Open Source Software.* 2018. **3.** 1026. doi:10.21105/joss.01026.

10. Abid, A. *et al. Gradio: Hassle-Free Sharing and Testing of ML Models in the Wild* in *ICML : Workshop on Human In the Loop Learning (HILL)* Publisher: arXiv Version Number: 1 (Long Beach, California, USA, 2019). https://arxiv.org/abs/1906.02569.

11. Jovic, A., Brkic, K. & Bogunovic, N. *A review of feature selection methods with applications* in *2015 38th International Convention on Information and Communication Technology, Electronics and Microelectronics (MIPRO)* 2015 38th International Convention on Information and Communication Technology, Electronics and Microelectronics (MIPRO) (IEEE, Opatija, Croatia, May 2015), 1200–1205. ISBN: 978-953-233-082-3. http://ieeexplore.ieee.org/document/7160458/.

12. Lundberg, S. & Lee, S.-I. *A Unified Approach to Interpreting Model Predictions* in *Proceedings of the 31st International Conference on Neural Information Processing Systems* (2017). arXiv: 1705.07874[cs, stat]. http://arxiv.org/abs/1705.07874.

13. Woods, D. L. & Martin, J. L. Cortisol and Wake Time in Nursing Home Residents With Behavioral

Symptoms of Dementia. *Biological Research For Nursing*. 2007. **9.** 21–29. doi:10.1177/1099800407303982.

14. Gehrman, P. R. *et al.* Sleep-Disordered Breathing and Agitation in Institutionalized Adults With Alzheimer Disease. *The American Journal of Geriatric Psychiatry.* 2003. **11.** 426–433. doi:10.1097/00019442-200307000-00005.

15. Shannon, C. E. A Mathematical Theory of Communication. *Bell System Technical Journal.* 1948. **27.** 379–423. doi:10.1002/j.1538-7305.1948.tb01338.x.

16. Kenji, N. & Neveen, H. *Assessment Of Daylight In Relation To The Agitation Levels Of People With Dementia* in *BSO Conference* Proceedings of BSO Conference 2016: Third Conference of IBPSA-England. **3.** Journal Abbreviation: BSO Conference (IBPSA-England, Newcastle, UK, Sept. 2016), 170–177. https://publications.ibpsa.org/conference/paper/?id=bso2016_1013.
